# The National COVID Cohort Collaborative: Clinical Characterization and Early Severity Prediction

**DOI:** 10.1101/2021.01.12.21249511

**Authors:** Tellen D. Bennett, Richard A. Moffitt, Janos G. Hajagos, Benjamin Amor, Adit Anand, Mark M. Bissell, Katie Rebecca Bradwell, Carolyn Bremer, James Brian Byrd, Alina Denham, Peter E. DeWitt, Davera Gabriel, Brian T. Garibaldi, Andrew T. Girvin, Justin Guinney, Elaine L. Hill, Stephanie S. Hong, Hunter Jimenez, Ramakanth Kavuluru, Kristin Kostka, Harold P. Lehmann, Eli Levitt, Sandeep K. Mallipattu, Amin Manna, Julie A. McMurry, Michele Morris, John Muschelli, Andrew J. Neumann, Matvey B. Palchuk, Emily R. Pfaff, Zhenglong Qian, Nabeel Qureshi, Seth Russell, Heidi Spratt, Anita Walden, Andrew E. Williams, Jacob T. Wooldridge, Yun Jae Yoo, Xiaohan Tanner Zhang, Richard L. Zhu, Christopher P. Austin, Joel H. Saltz, Ken R. Gersing, Melissa A. Haendel, Christopher G. Chute, N3C Consortium

**Affiliations:** Section of Informatics and Data Science, Department of Pediatrics, University of Colorado School of Medicine, University of Colorado, Aurora, CO, USA; Department of Biomedical Informatics, Stony Brook University, Stony Brook, NY, USA; Stony Brook University, Stony Brook, NY, USA; Palantir Technologies, Denver, CO, USA; The University of Michigan at Ann Arbor, Ann Arbor, MI, USA; University of Rochester Medical Center, Rochester, NY, USA; Johns Hopkins University School of Medicine, Baltimore, MD, USA; Sage Bionetworks, Seattle, WA, USA; University of Kentucky, Lexington, KY, USA; Real World Solutions, IQVIA, Cambridge, MA, USA; Observational Health Data Sciences and Informatics, New York, NY, USA; University of Alabama at Birmingham, Birmingham, AL, USA; Translational and Integrative Sciences Center, Oregon State University, Corvallis, OR, USA; Department of Biomedical Informatics, University of Pittsburgh, Pittsburgh, PA, USA; TriNetX, Cambridge, MA, USA; North Carolina Translational and Clinical Sciences Institute (NC TraCS), University of North Carolina at Chapel Hill, Chapel Hill, NC, USA; University of Texas Medical Branch, Galveston, TX, USA; Oregon Clinical and Translational Research Institute, Oregon Health & Science University, Portland, OR, USA; Tufts Medical Center Clinical and Translational Science Institute, Tufts Medical Center, Boston, MA, USA; National Center for Advancing Translational Sciences, National Institutes of Health, Bethesda, MD, USA; Translational and Integrative Sciences Center, Dept. of Molecular Toxicology, Oregon State University, Corvallis, OR, USA; Schools of Medicine, Public Health, and Nursing, Johns Hopkins University, Baltimore, MD, USA

## Abstract

**Background:** The majority of U.S. reports of COVID-19 clinical characteristics, disease course, and treatments are from single health systems or focused on one domain. Here we report the creation of the National COVID Cohort Collaborative (N3C), a centralized, harmonized, high-granularity electronic health record repository that is the largest, most representative U.S. cohort of COVID-19 cases and controls to date. This multi-center dataset supports robust evidence-based development of predictive and diagnostic tools and informs critical care and policy.

**Methods and Findings:** In a retrospective cohort study of 1,926,526 patients from 34 medical centers nationwide, we stratified patients using a World Health Organization COVID-19 severity scale and demographics; we then evaluated differences between groups over time using multivariable logistic regression. We established vital signs and laboratory values among COVID-19 patients with different severities, providing the foundation for predictive analytics. The cohort included 174,568 adults with severe acute respiratory syndrome associated with SARS-CoV-2 (PCR >99% or antigen <1%) as well as 1,133,848 adult patients that served as lab-negative controls. Among 32,472 hospitalized patients, mortality was 11.6% overall and decreased from 16.4% in March/April 2020 to 8.6% in September/October 2020 (p = 0.002 monthly trend). In a multivariable logistic regression model, age, male sex, liver disease, dementia, African-American and Asian race, and obesity were independently associated with higher clinical severity. To demonstrate the utility of the N3C cohort for analytics, we used machine learning (ML) to predict clinical severity and risk factors over time. Using 64 inputs available on the first hospital day, we predicted a severe clinical course (death, discharge to hospice, invasive ventilation, or extracorporeal membrane oxygenation) using random forest and XGBoost models (AUROC 0.86 and 0.87 respectively) that were stable over time. The most powerful predictors in these models are patient age and widely available vital sign and laboratory values. The established expected trajectories for many vital signs and laboratory values among patients with different clinical severities validates observations from smaller studies, and provides comprehensive insight into COVID-19 characterization in U.S. patients.

**Conclusions:** This is the first description of an ongoing longitudinal observational study of patients seen in diverse clinical settings and geographical regions and is the largest COVID-19 cohort in the United States. Such data are the foundation for ML models that can be the basis for generalizable clinical decision support tools. The N3C Data Enclave is unique in providing transparent, reproducible, easily shared, versioned, and fully auditable data and analytic provenance for national-scale patient-level EHR data. The N3C is built for intensive ML analyses by academic, industry, and citizen scientists internationally. Many observational correlations can inform trial designs and care guidelines for this new disease.

## Introduction

As of mid-December 2020, severe acute respiratory syndrome associated with coronavirus-2 (SARS-CoV-2) has infected more than 70 million people and caused more than 1.6 million deaths worldwide^[a]^. SARS-CoV-2 can cause coronavirus disease of 2019 (COVID-19), a condition characterized by pneumonia, hyperinflammation, hypoxemic respiratory failure, a prothrombotic state, cardiac dysfunction, substantial mortality, and persistent morbidity in some survivors. Few FDA-authorized therapeutics are available, and vaccine deployment has been slow. Progress in COVID-19 research has been slowed by lack of broad access to clinical data. Investigators in the United Kingdom^1^ and Denmark^1,2^ have performed person-level analytics across their populace, but the U.S. has not had this capacity. A large, multi-center, representative clinical dataset has been desperately needed by U.S. clinicians, scientists, health systems, and policy-makers to develop predictive and diagnostic computational tools and to inform critical decisions.

To address these gaps, N3C was formed to accelerate understanding of SARS-CoV-2 and demonstrate a novel approach for collaborative data sharing and analytics during a pandemic. The National COVID Cohort Collaborative (N3C)^3^ is comprised of members from the NIH Clinical and Translational Science Awards (CTSA) Program and its Center for Data to Health (CD2H), the IDeA Centers for Translational Research^[b]^, the National Patient-Centered Clinical Research Network (PCORNet, pcornet.org), the Observational Health Data Sciences and Informatics (OHDSI, ohdsi.org) network, TriNetX (trinetx.com), and the Accrual to Clinical Trials (ACT, actnetwork.us/National) network.

Here we provide a detailed clinical description of the largest cohort of U.S. COVID-19 cases and representative controls to date. This cohort is racially and ethnically diverse and geographically distributed. We demonstrate its impact by 1) evaluating COVID-19 severity and risk factors over time and 2) using machine learning (ML) to develop a clinically useful model that accurately predicts severity using data from the first day of hospital admission.

## Methods

### Cohort Definition and Outcome Stratification

Because of the broad inclusion criteria, N3C includes cases and appropriate controls for varied analyses including both outpatients and inpatients (Supplemental Table 1). N3C includes patients with any encounter after 1/1/2020 with 1) one of a set of *a priori*-defined SARS-CoV-2 laboratory tests or 2) a “strong positive” diagnostic code or 3) two “weak positive” diagnostic codes during the same encounter or on the same date prior to 5/1/2020. The cohort definition is publicly available on GitHub.^[c]^ For N3C patients, encounters in the same health system beginning on or after 1/1/2018 are also included to provide information about pre-existing health conditions (“lookback data”). See Supplemental Methods for information about N3C architecture, data ingestion, and integration.

We conducted a retrospective cohort study of adults ≥ 18 years old at the 34 N3C sites whose data 1) have completed harmonization and integration (see Supplemental Methods), 2) were released for analysis, and 3) included the necessary death and mechanical ventilation information (Supplemental Figure 1). In order to demonstrate the scope of N3C, Figure 1a-b and Supplemental Table 1 are based on the entire cohort. All subsequent analyses include only patients with a positive SARS-CoV-2 laboratory test (polymerase chain reaction [PCR] or antigen) (Table 1).

**Figure 1:**
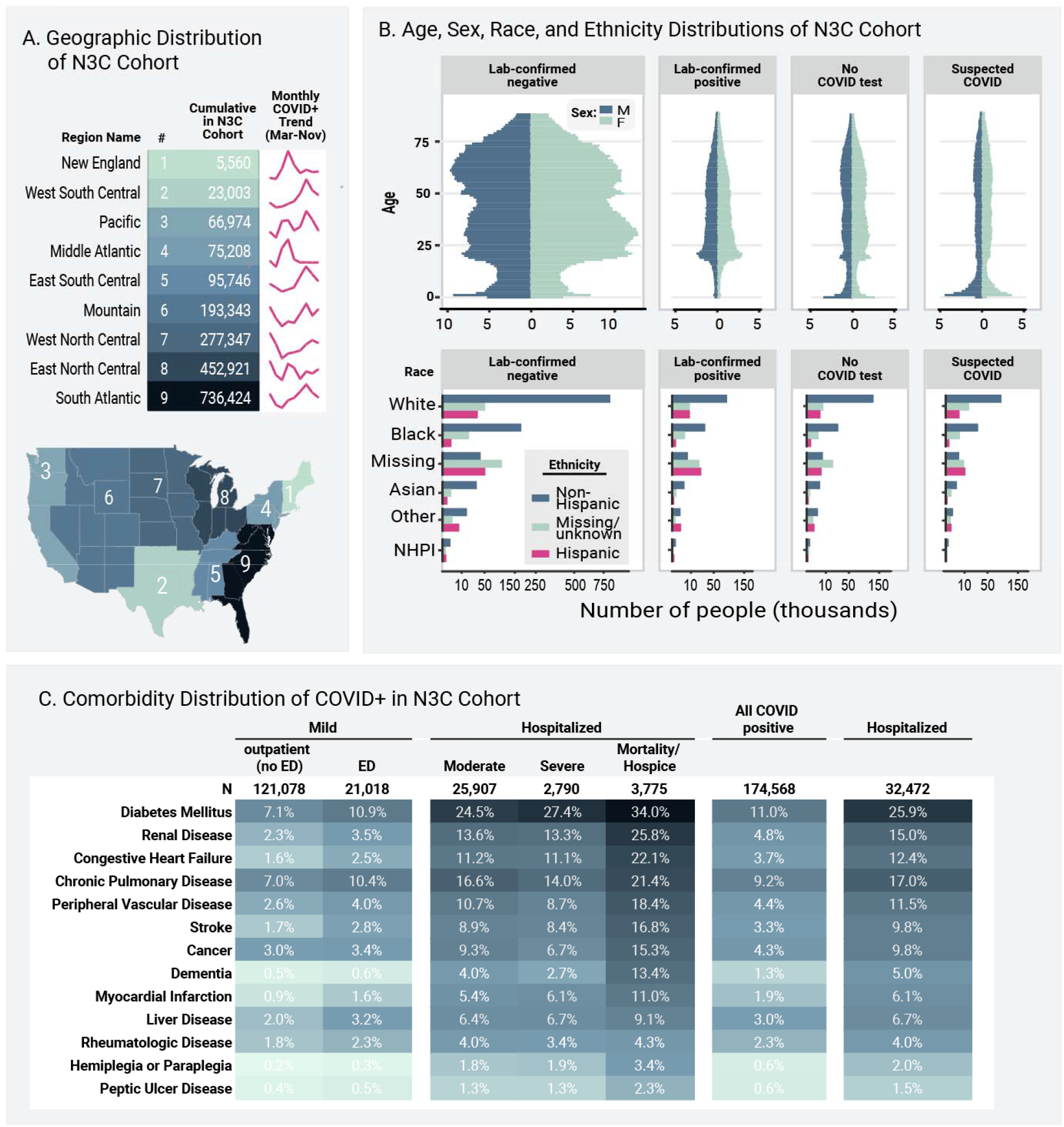
Geographic, Age, Sex, Race, Ethnicity, and Comorbidity Distributions of N3C Cohort. **Figure 1a** shows the representation of each U.S. subregion in the overall (N = 1,926,526) cohort. Trend lines show the accumulation of each subregion’s sample size of lab confirmed positive cases over 2020. The Southeast, Mid-Atlantic, and Midwestern regions are the most heavily represented, but all regions have substantial patient counts. **Figure 1b** shows the age, sex, race, and ethnicity distributions of the overall N3C cohort, stratified by the N3C phenotype groups (publicly available on GitHub^[c]^). Racial and ethnic minorities are well-represented. COVID = coronavirus disease. NHPI = Native Hawaiian or Pacific Islander. **Figure 1c** shows comorbidity distributions for the laboratory-confirmed positive adult cohort (N = 174,568). See Supplemental Methods for comorbidity definitions. We stratified patients using the Clinical Progression Scale (CPS) established by the World Health Organization (WHO) for COVID-19 clinical research, see Table 1^4^. Severity assigned by patient-specific encounter maximum severity. No ED = outpatient only without emergency department visit, ED = emergency department visit, moderate = hospitalized without invasive ventilation or extracorporeal membrane oxygenation (ECMO), severe = hospitalized with invasive ventilation or ECMO, mortality/hospice = hospital mortality or discharge to hospice.

**Table 1:** SARS-CoV-2 Laboratory-Confirmed Positive Cohort Characteristics and Clinical Course. **Table 1 Legend**. SARS-CoV-2 = severe acute respiratory syndrome associated with coronavirus-2. ED = Emergency Department. WHO = World Health Organization. ECMO = extracorporeal membrane oxygenation. LOS = length of stay. We stratified patients using the Clinical Progression Scale (CPS) established by the World Health Organization (WHO) for COVID-19 clinical research.^4^ Severity assigned by patient-specific encounter maximum severity. *Other includes non-binary, no matching concept, and no information. Per N3C policy, we censored any cells with 1-20 patients and replaced them with “20 or fewer.”

|  | Mild<br>Outpatient<br>WHO Severity 1-3 | Mild_ED<br>Outpatient with<br>ED visit<br>WHO Severity<br>~3 | Moderate<br>Hospitalized<br>without invasive<br>ventilation<br>WHO<br>Severity 4-6 | Severe<br>Hospitalized with<br>invasive<br>ventilation or<br>ECMO<br>WHO<br>Severity 7-9 | Hospital<br>Mortality or<br>Discharge to<br>Hospice<br>WHO Severity<br>10 |
| --- | --- | --- | --- | --- | --- |
| **N** | 121,078 | 21,018 | 25,907 | 2,790 | 3,775 |
| **Age (mean +/- SD)** | 41.1 (17.2)<br>n=121078 | 43.4 (16.8)<br>n=21018 | 55.0 (19.1)<br>n=25907 | 57.0 (15.4)<br>n=2790 | 71.8 (14.7)<br>n=3775 |
| **Sex** |  |  |  |  |  |
| **Female** | 65,435 | 11,410 | 13,396 | 1,089 | 1,564 |
| **Male** | 55,526 | 9,605 | 12,506 | 1,697 | 2,211 |
| **Other\*** | 117 | 20 or fewer | 20 or fewer | 20 or fewer | 0 |
| **Race** |  |  |  |  |  |
| **White or Caucasian** | 70,330 | 7,786 | 10,739 | 1,020 | 1,912 |
| **Black or African-American** | 14,616 | 6,351 | 8,003 | 869 | 1,101 |
| **Native Hawaiian or Pacific Islander** | 267 | 40 | 66 | 20 or fewer | 20 or fewer |
| **Asian** | 2,778 | 564 | 717 | 86 | 120 |
| **Other** | 1030 | 403 | 373 | 51 | 48 |
| **Missing/Unknown** | 32,057 | 5,874 | 6,009 | 757 | 584 |
| **Ethnicity** |  |  |  |  |  |
| **Hispanic** | 18,539 | 5,312 | 5,145 | 610 | 476 |
| **Non-Hispanic** | 80,188 | 12,510 | 17,313 | 1,789 | 2,779 |
| **Missing/Unknown** | 22,351 | 3,196 | 3,449 | 391 | 520 |
| **Insurance Payer** |  |  |  |  |  |
| **Medicare** | 2,480 | 906 | 2,852 | 308 | 823 |
| **Commercial** | 11,718 | 2,277 | 1,984 | 227 | 237 |
| **Medicaid** | 2,945 | 1,590 | 1,974 | 242 | 294 |
| **Other** | 115,480 | 18,576 | 22,876 | 2,409 | 3,124 |
| **Body Size** |  |  |  |  |  |
| **Body Mass Index**<br>**(mean +/- SD)** | 30.1 (7.6)<br>n=39836 | 31.2 (7.8) n=9552 | 31.0 (9.0)<br>n=16489 | 32.9 (9.4) n=1862 | 29.5 (8.7) n=2440 |
| **Weight, kg**<br>**(mean +/- SD)** | 86.3 (23.7)<br>n=47284 | 87.3 (23.7)<br>n=13511 | 88.6 (26.0)<br>n=20068 | 95.5 (26.8)<br>n=2349 | 84.6 (26.7)<br>n=3106 |
| **Clinical course** |  |  |  |  |  |
| **Hospital LOS, median (IQR)** |  |  | 6.6 (8.9) n=25906 | 27.5 (26.1)<br>n=2790 | 14.0 (23.3)<br>n=3775 |

### Hospital Index Encounter and Clinical Severity

We defined a single index encounter for each laboratory-confirmed positive patient using a pre-specified algorithm (Supplemental Methods). We stratified patients using the Clinical Progression Scale (CPS) established by the World Health Organization (WHO) for COVID-19 clinical research.^4^ We placed patients into strata defined by the maximum clinical severity during their index encounter (Table 1). We collapsed some WHO CPS categories due to data limitations (e.g. some sites do not submit fraction of inspired oxygen [FiO_2_]).

### Variable Definition and Statistical Methods

We defined or identified existing concept sets in the Observational Medical Outcomes Partnership (OMOP) common data model (CDM) for each clinical concept (e.g. laboratory measure, vital sign, or medication, see Supplemental Methods). We validated each concept set with input from informatics and clinical subject matter experts. All concept sets and analytic pipelines are fully reproducible and will be made publicly available. We tested time trends using linear regression and differences between groups using multivariable logistic regression. See Supplemental Methods for additional information including software packages used.

### Machine Learning Methods

We developed models to predict patient-specific maximum clinical severity: hospitalization with death, discharge to hospice, invasive mechanical ventilation, or extracorporeal membrane oxygenation (ECMO) versus hospitalization without any of those. To avoid immortal time bias, we only included patients with at least one hospital overnight. We split the hospitalized laboratory-confirmed positive cohort into randomly selected 70% training and 30% testing cohorts stratified by outcome proportions and held out the testing set. We chose a broad set of potential predictors present for at least 15% of the training set (Supplemental Table 2). The input variables are the most abnormal value on the first calendar day of the hospital encounter. When patients did not have a laboratory test value on the first calendar day, we imputed normal values for specialized labs (e.g. ferritin, procalcitonin) and the median cohort value for common labs (e.g. sodium, albumin) (Supplemental Table 2). We compared several analytical approaches with varying flexibility and interpretability: logistic regression +/- L1 and L2 penalty, random forest, support vector machines, and XGBoost (github.com/dmlc/xgboost).

We internally validated models and limited overfitting using 5-fold cross-validation and evaluated models using the testing set and area under the receiver operator characteristic (AUROC) as the primary metric. Secondary metrics included precision/positive predictive value, recall/sensitivity, specificity, and F1-measure. Because SARS-CoV-2 outcomes have improved over time^5^, we evaluated model performance overall and for March-May 2020 and June-October 2020. See Supplemental Methods.

## Results

### Study Cohort

As of December 7, 2020, data from 34 sites was harmonized and integrated into the N3C release set. The cohort includes data about 1,926,526 patients (Supplemental Table 1). The cohort derives from all U.S. geographic regions, but is more concentrated in the Southeast, Mid-Atlantic, and Midwest (Figure 1a). The age, sex, race, ethnicity, and insurance payer distributions (Figure 1b and Supplemental Table 1) indicate a diverse patient cohort that is representative of many segments of the U.S. population. Importantly, African-American and Hispanic patients, who have suffered disproportionately from COVID-19^6^, are represented in sufficient numbers to support robust subgroup analyses, pathophysiologic hypothesis generation, and testing of algorithms and models to avoid bias (Table 1). Supplemental Tables 3a and 3b show the cohort stratified by CDM and strengths and weaknesses of each CDM. Figure 1a shows cohort geographic distribution evolution during 2020.

Of the overall cohort, 174,568 adults (9.1%) had a positive SARS-CoV-2 PCR or antigen test at a site with death and ventilation data available (Table 1). Antigen tests represent <5% of a single site’s positive tests. All other positive patients had positive PCR tests.

### Clinical Course and Mortality

Of those with a positive test, 32,472 (18.6%) were hospitalized. The median length of hospital stay was 5 days (IQR 2 to 10). Mortality (including discharge to hospice) was 11.6% among hospitalized patients (Table 1). Others have reported that inpatient mortality has decreased over time^7^. We confirm this: inpatient mortality decreased from 16.4% in March and April to 8.6% in September and October (P for monthly linear trend 0.002). Our data also show that clinical severity has shifted toward less invasive mechanical ventilation and/or ECMO as the pandemic has progressed (Figure 2a).

**Figure 2.**
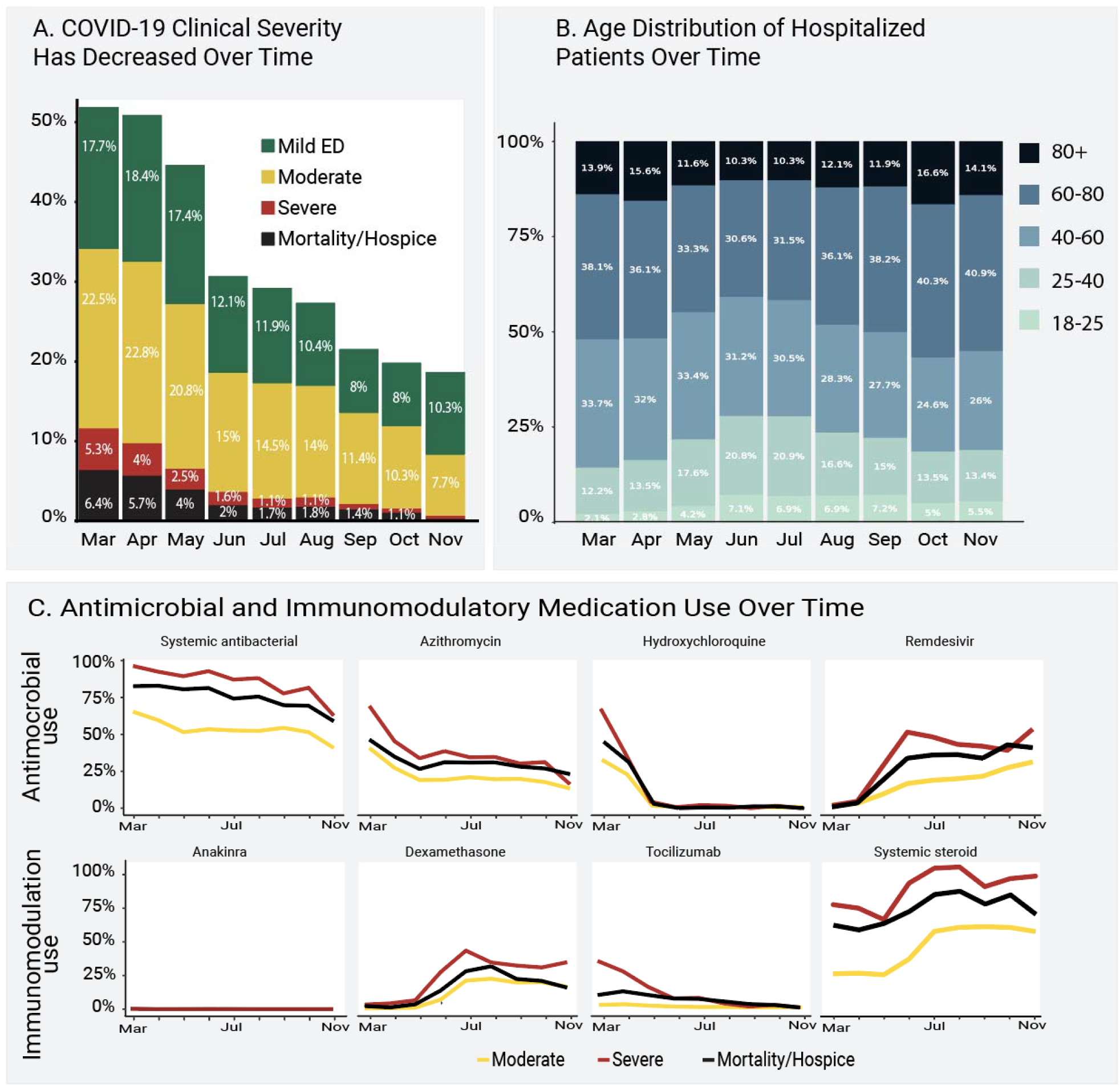
Clinical Severity, Age, and Antimicrobial and Immunomodulatory Medication Use Over Time. **Figure 2a** shows the distribution of patient-specific encounter maximum severity among hospitalized patients during 2020. Mortality and invasive ventilation or extracorporeal membrane oxygenation (“Severe”) have decreased steadily, monthly trend p = 0.002. Strata assigned using the Clinical Progression Scale (CPS) established by the World Health Organization (WHO) for COVID-19 clinical research (hospital mortality or discharge to hospice [black], invasive ventilation or extracorporeal membrane oxygenation [red], hospitalized without any of those [yellow], or emergency department visit only [green], see Table 1^4^). The percentage of patients from each month is shown over each severity group bar. **Figure 2b** shows how the age distribution of hospitalized patients has changed during 2020. The percentage of patients from each month is shown over each age bracket bar. Older patients (darker blue) were more prominent in the spring and the fall, with more younger patients (lighter blue/teal) in the summer. **Figure 2c** shows the evolution of antimicrobial and immunomodulatory treatment regimens for hospitalized patients (top 3 severity strata, see Table 1) during 2020.

### Demographics, Comorbidities, and Obesity

The age distribution for hospitalized patients was older during spring 2020, younger during the summer, and older again in the fall (Figure 2b). Lookback data that allowed calculation of comorbidities was present for 49% of hospitalized patients. Of hospitalized patients, 41% had at least one comorbid condition; the most common was diabetes mellitus (25.9%, Figure 1c). Mean body mass index (BMI) was 30 or above for all severity groups (Table 1). In a multivariable logistic regression model, age, male sex, liver disease, dementia, African-American and Asian race, and obesity (BMI > 30) were independently associated with higher patient-specific maximum clinical severity (invasive ventilation, ECMO, death, or discharge to hospice versus none of those, Supplemental Table 4). Interestingly, rheumatologic disease and blood type AB were protective. This analysis was conducted only to provide inference about previously reported risk factors and occurred after the prediction model was built, see below.

### Vital Sign and Laboratory Measurements

As a hospital encounter progressed, those who ultimately developed higher clinical severity (invasive ventilation, ECMO, or death) tended to have progressively more abnormal (higher) mean heart rate (HR), respiratory rate (RR), and temperature than those who did not (Figure 3a). Mean diastolic blood pressure (DBP) and oxygen saturation (SpO_2_) among those who ultimately died continued to become more abnormal (lower) while those who were invasively ventilated or on ECMO became more normal (higher, Figure 3a). Early in the hospital encounter, mean values of DBP, SpO_2_, and widely used measures of inflammation (C-reactive protein [CRP] and ferritin), immunologic activation (white blood cell count, WBC), fibrinolysis (D-dimer), oxygen delivery (lactate), and renal function (creatinine) were more abnormal among those who ultimately required invasive ventilation or ECMO than those who did not (Figures 3a and 3b). These findings support the hypothesis that clinical severity can be predicted using information available early in a hospital course (see prediction models).

**Figure 3.**
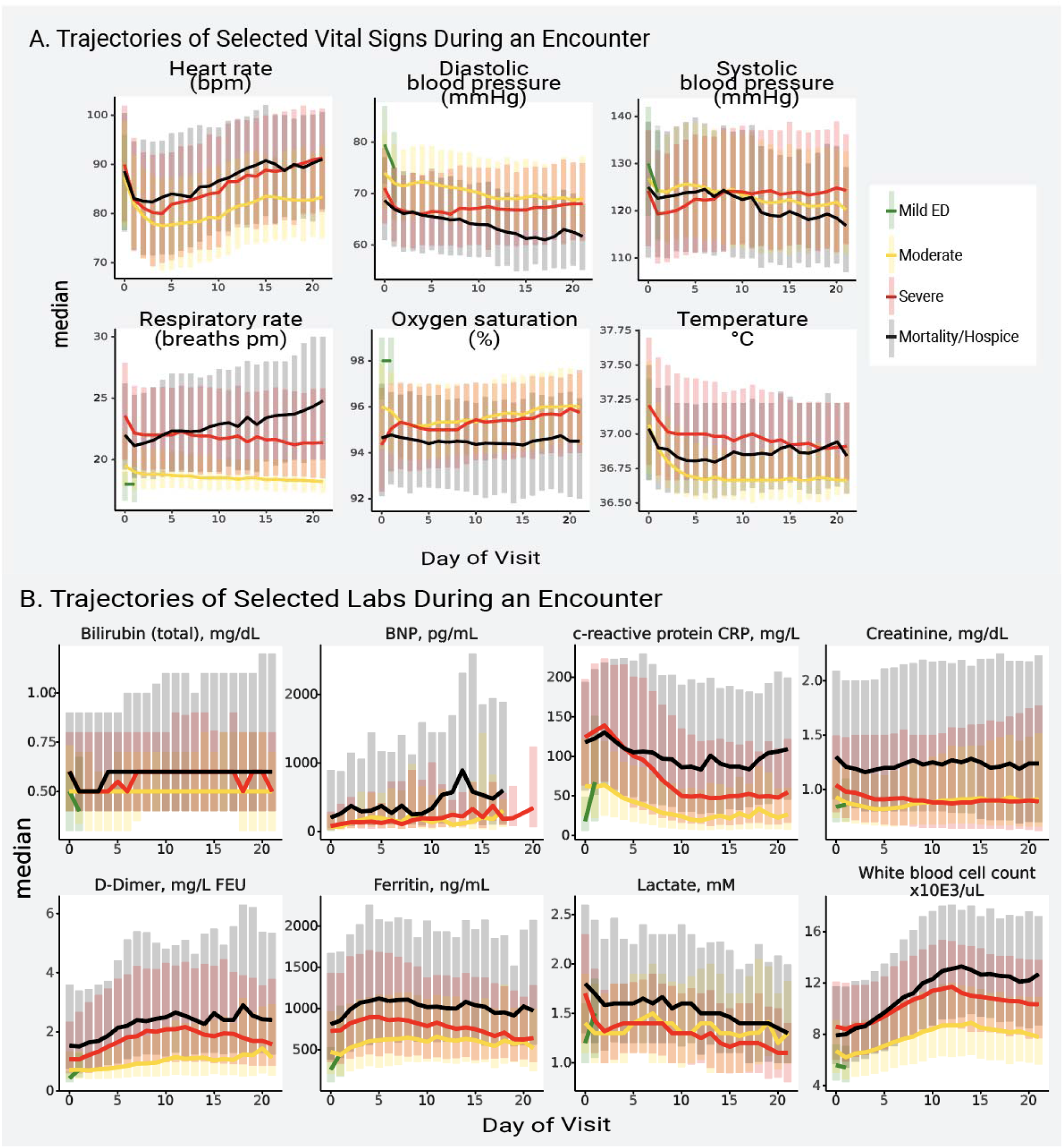
Trajectories of Vital Signs and Laboratory Tests During a Hospital Encounter. Figure 3a shows the median (line) and interquartile range (bars) of each vital sign on each hospital day, stratified by patient maximum severity (hospital mortality or discharge to hospice [black], invasive ventilation or extracorporeal membrane oxygenation [red], hospitalized without any of those [yellow], or emergency department visit only [green], see Table 1). Figure 3b shows the median (line) and interquartile range (bars) of each laboratory test on each hospital day, stratified by the same severity groups. BNP = brain natriuretic peptide.

Other measurements (e.g. sodium, platelet count, lymphocyte count) show potential utility as early outcome predictors, as their values near the beginning of a hospital encounter tend to separate patients with lower and higher maximum clinical severity (Supplemental Figure 2). Mean values of brain natriuretic peptide were low early in hospital encounters but showed meaningful spikes between hospital days 10 and 15. This is consistent with reports of the timing of cardiac failure in COVID-19^8^. Overall, patients with more abnormal nadir and/or peak values of several vital signs and laboratory measurements were more often represented in higher severity groups (invasively ventilated, ECMO, or death; Supplemental Figures 3a-b). CRP, ferritin, D-dimer, WBC, and IL-6 have been identified by the WHO as key biochemical parameters for a core COVID-19 outcome set^4^. These were measured in 44-94% of hospitalized patients, except IL-6 (7.6%). A relatively small number of hospitalized patients had blood type data (9.1%, Supplemental Figure 4).

### Treatments

Usage of antimicrobial and immunomodulatory medications has changed dramatically over time (Figure 2c). Overall, 66.2% of the hospitalized cohort received at least one antimicrobial, with significant treatment regimen heterogeneity (Supplemental Figure 5a and Supplemental Table 5a). Patients who received invasive ventilation and ECMO received more antimicrobials overall (Supplemental Figure 5a). Antivirals with potential activity against SARS-CoV-2 were given to 16.7% (remdesivir) and 0.6% (lopinavir/ritonavir) of hospitalized patients. At least one immunomodulatory medication was given to 41.5% of hospitalized patients, also with wide variation in treatment regimen (Supplemental Figure 5b and Supplemental Table 5a). More patients received hydrocortisone, methylprednisolone, and prednisone than dexamethasone (Supplemental Table 5a). The trial indicating survival benefit from dexamethasone was published in July 2020.^9^ Other steroids also have modestly supportive clinical trial data.^10^

Of the hospitalized cohort, 14.0% received any invasive respiratory support (mechanical ventilation or inhaled or systemic pulmonary vasodilators, Supplemental Table 5b). Similarly, 8.3% received medications for cardiovascular support or ECMO and 3.2% received dialysis or continuous renal replacement therapy.

### Severity Prediction

We developed several models that accurately predict a severe clinical course using data from the first hospital calendar day (Supplemental Figure 6 and Supplemental Table 6). The models with the best discrimination of severe versus non-severe clinical course were built using XGBoost (AUROC 0.87) and random forest (AUROC 0.86). Both are flexible nonlinear tree-based models that provide interpretability with a variable importance metric (Figure 4). Importantly, discrimination by the two models was stable over time (March-May 2020 and June-October 2020, Supplemental Table 6). This indicates that the models did not train on health care processes only typical during the pandemic’s chaotic first wave. Commonly collected variables (age, SpO_2_, RR, blood urea nitrogen, systolic blood pressure, and aspartate aminotransferase) were among the inputs with the highest variable importance for both models (Figure 4).

**Figure 4.**
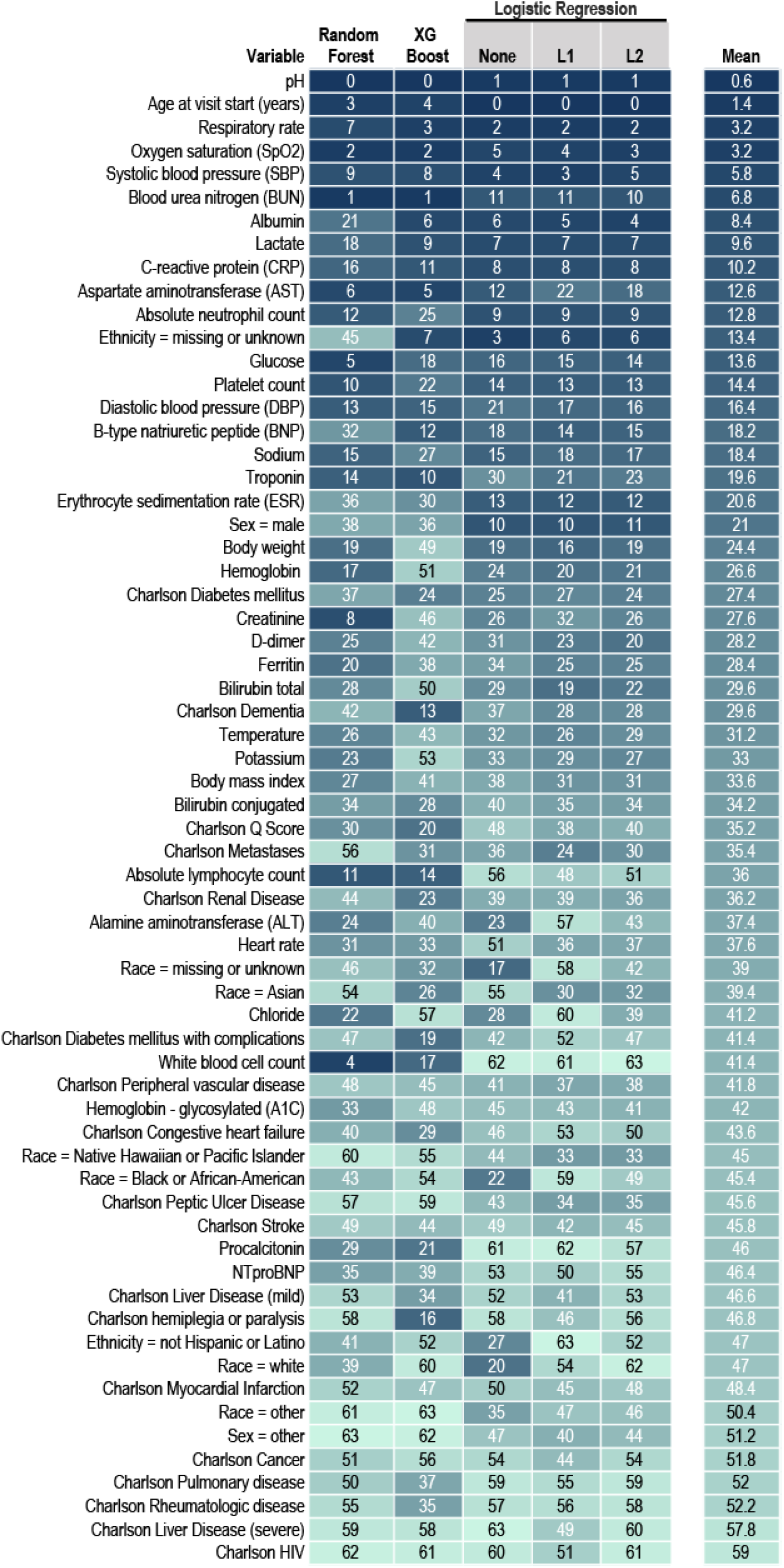
Variable Importance in the Machine Learning Models Predicting Clinical Severity. The 64 machine learning (ML) model input variables are listed by their mean variable importance rank across ML model types. Each column is a ML model type. Logistic regression is shown without penalization and with L1 and L2 penalties. The table cells show a heat map with darkest (blue) representing highest variable importance and lightest (teal) representing lower variable importance. See Methods and Supplemental Methods for details about variable definitions, model construction, and testing. NTproBNP = N-Terminal-prohormone B-type Natriuretic Peptide.

## Discussion

This manuscript characterizes the largest U.S. COVID-19 cohort to date. We have confirmed a month-over-month decrease in COVID-19 inpatient mortality and invasive ventilation rates since March 2020. We developed accurate ML models to predict clinical severity based only on information available on the first calendar day of admission. The most powerful predictors in these models are patient age and widely available vital sign and laboratory values. These models can be the basis for generalizable clinical decision support tools. We also established expected trajectories for many vital signs and laboratory values among patients with different clinical severities. Expected trajectories can contribute to clinician decision-making about what a patient will need.

Site heterogeneity in the distribution of predictors of severe COVID-19 disease including age, race, ethnicity, and existing comorbidities (e.g. diabetes) has complicated interpretation of their independent impact on outcomes. Like others, we found that age, male sex^1^, African-American race^6,11^and obesity^12,13^ were associated with greater clinical severity. Associations of liver disease and dementia with COVID-19 severity have also been reported^14,15^. We found that patients with rheumatologic disease had lower clinical severity. This is consistent with reports that after adjustment for age, diabetes, and renal impairment, patients with rheumatologic disease on some treatment regimens have lower risk of hospitalization^16^. Increased risk of intubation and death has been inconsistently found among patients with blood types AB, A, and B relative to type O.^17–19^ In contrast, we found that blood type AB was protective.

We also found significant treatment regimen heterogeneity for inpatients with COVID-19. Some medications have fallen out of favor (e.g. hydroxychloroquine, azithromycin); others are the subject of ongoing studies (e.g. anakinra, tocilizumab). For most treatments, the balance of risks and benefits has not been evaluated rigorously in randomized controlled trials. Ongoing monitoring for adverse effects in observational data like N3C will be important.

The N3C has unique features that distinguish it from other COVID-19 data resources. First, it harmonizes data from a very large number of clinical sites (73 have signed data transfer agreements to date). This is important because significant site-level variation in critical metrics such as invasive ventilation and mortality has been reported.^20–23^ Central curation ensures that N3C data are robust and quality-assured across sites. This is in contrast to the known challenges of relying on site-level CDM quality assurance processes in distributed networks (e.g. OHDSI, PCORnet). Most U.S. reports of COVID-19 clinical characteristics, disease course, treatments, and outcomes come from a single hospital or health system^6,22^ in a single geographic region. Another network has reported a large COVID-19 cohort, but the patient-level data is not centralized and thus is less amenable to machine learning^24^.

Developed under the intense time pressure of a health crisis, earlier data aggregation efforts ^1,21,25–28^ may not have been designed to support future research. The N3C Data Enclave^3^ provides transparent, easily shared, versioned, and fully auditable data and analytic provenance. This is a key advantage, as a lack of auditable data and analytic provenance has resulted in retraction of high-profile COVID-19 publications.^29,30^

N3C users should bear in mind its limitations. Because the data are aggregated from many health systems and 4 CDMs that vary in granularity, some sites have systematic missingness of some variables (see Supplemental Methods). Detailed respiratory support information such as oxygen flow, FiO_2_, and ventilator settings (typically recorded in EHR flowsheets) is not fully available. Orders related to limitations in care such as “do not attempt resuscitation” (DNAR) are not yet present in N3C. Some inpatient mortality in our study likely occurred in patients who had DNAR orders in place. Exclusion of those patients might improve severity model prediction. Finally, exact time of laboratory values is inconsistently provided by sites, so labs are standardized to calendar day, but not time of day.

In conclusion, N3C is a nationally representative, transparent, reproducible, harmonized data resource that enables effective and efficient collaborative observational COVID-19 research. N3C is built for intensive machine learning analyses by academic, industry, and citizen scientists internationally. We have demonstrated its utility by developing a clinically useful patient severity predictor.

### Ethics and Regulatory

The N3C data transfer to NCATS is performed under a Johns Hopkins University Reliance Protocol # IRB00249128 or individual site agreements with NIH.

Use of the N3C data for this study is authorized under the following IRB Protocols:

| **Site** | **IRB name** | **Exempted vs. approved** | **Protocol number** |
| --- | --- | --- | --- |
| University of Alabama-Birmingham | The University of Alabama at Birmingham Office of the Institutional Review Board for Human Use | exempted | IRB-300006285 |
| University of Colorado | Colorado Multiple Institutional Review Board | approved | 20-2225 |
| Johns Hopkins University | Johns Hopkins Office of Human Subjects Research - Institutional Review Board | approved | IRB00249128 |
| University of Kentucky | Medical Institutional Review Board of the University of Kentucky | exempted | 62294 |
| University of Michigan | University of Michigan Medical School Institutional Review Board | approved | HUM00188854 |
| University of North Carolina | University of North Carolina Chapel Hill Institutional Review Board | exempted | 20-3106 |
| Oregon State University | Oregon State University Institutional Review Board | approved | IRB-2020-0830 |
| University of Rochester | University of Rochester Research Subjects Review Board | exempted | STUDY00005366 |
| Stony Brook University | Office of Research Compliance, Division of Human Subject Protections, Stony Brook University | exempted | IRB2020-00604 |
| University of Texas-Medical Branch | Institutional Review Board of the University of Texas Medical Branch | exempted | 20-0245 |

The N3C Data Enclave is approved under the authority of the NIH Institutional Review Board for Protocol 000082 associated with NIH iRIS reference number: 546652 entitled: “NCATS National COVID-19 Cohort Collaborative (N3C) Data Enclave Repository.” Further information can be found at https://ncats.nih.gov/n3c/resources.

## Data Availability

The N3C Data Enclave (covid.cd2h.org/enclave) houses fully reproducible, transparent, and broadly available limited and de-identified datasets (HIPAA definitions: https://www.hhs.gov/hipaa/for-professionals/privacy/special-topics/de-identification/index.html). Data is accessible by investigators at institutions that have signed a Data Use Agreement with NIH who have taken human subjects and security training and attest to the N3C User Code of Conduct. Investigators wishing to access the limited dataset must also supply an institutional IRB protocol. All requests for data access are reviewed by the NIH Data Access Committee. A full description of the N3C Enclave governance has been published;3 information about how to apply for access is available on the NCATS website: https://ncats.nih.gov/n3c/about/applying-for-access. Reviewers and health authorities will be given access permission and guidance to aid reproducibility and outcomes assessment. A Frequently Asked Questions about the data and access has been created at; https://ncats.nih.gov/n3c/about/program-faq
The data model is OMOP 5.3.1, specifications are posted at: https://ncats.nih.gov/files/OMOP_CDM_COVID.pdf
The latest version of the N3C Covid-19 Phenotype is always available at:
https://github.com/National-COVID-Cohort-Collaborative/Phenotype_Data_Acquisition
Governance documents, codesets, code, and other N3C resources are available within the project Github repositories and/or in Zenodo for archival purposes:
https://github.com/National-COVID-Cohort-Collaborative
https://zenodo.org/communities/cd2h-covid/
Information on the source Common Data Models is available at:
OHDSI: https://ohdsi.org/
PCORNet: https://pcornet.org/
ACT: https://www.dbmi.pitt.edu/node/53983
TriNetX: https://trinetx.com/
Other referenced resources are available at:
COVID-19 Map - Johns Hopkins Coronavirus Resource Center https://coronavirus.jhu.edu/map.html
Institutional Development Award Program Infrastructure for Clinical and Translational Research (IDeA-CTR) https://www.nigms.nih.gov/Research/DRCB/IDeA/Pages/IDeA-CTR.aspx
xgboost https://github.com/dmlc/xgboost

https://covid.cd2h.org/enclave

https://github.com/National-COVID-Cohort-Collaborative

https://zenodo.org/communities/cd2h-covid/

## Acknowledgments

The analyses described in this publication were conducted with data or tools accessed through the NCATS N3C Data Enclave covid.cd2h.org/enclave and supported by NCATS U24 TR002306 to Drs. Haendel, Guinney, Chute, Saltz, and Williams; also supporting Emily Pfaff, Anita Walden, Julie McMurry, Andrew Neumann, Davera Gabriel, and Harold Lehmann. Dr. Tellen D. Bennett was supported by UL1TR002535 03S2 and UL1TR002535; James Brian Byrd by NIH grant K23HL128909; Dr. Alina Denham by DP5OD021338 (PI: Hill); Dr. Ramakanth Kavuluru by NLM R01LM01324 and NCATS CTSA: UL1TR001998; Michele Morris by UL1TR001857-01S1 ACT; Seth Russell by the Data Science to Patient Value University of Colorado Anschutz Medical Campus; and Dr. Heidi Spratt by UL1TR001439. This research was possible because of the patients whose information is included within the data from participating organizations (covid.cd2h.org/dtas) and the organizations and scientists (covid.cd2h.org/duas) who have contributed to the on-going development of this community resource^3^.

Carilion Clinic (UL1TR003015-02S2: Provision of Clinical Data to Support a Nationwide COVID-19 Cohort Collaborative); George Washington Children’s Research Institute (UL1TR001876: Clinical and Translational Science Institute at Children’s National); Duke University (UL1TR002553: Duke CTSA); Johns Hopkins University (UL1TR003098: Johns Hopkins Institute for Clinical and Translational Research); Mayo Clinic Rochester (UL1TR002377: Mayo Clinic Center for Clinical and Translational Science); Medical University of South Carolina (UL1TR001450: South Carolina Clinical & Translational Research Institute SCTR); Penn State Health Milton S. Hershey Medical Center (UL1TR002014: Penn State Clinical and Translational Science Institute); Rush University Medical Center (UL1TR002389: Institute for Translational Medicine); Stony Brook University; The Ohio State University (UL1TR002733: The OSU Center for Clinical and Translational Science: Advancing Today’s Discoveries to Improve Health); Tufts University Boston (UL1TR002544-03S4: Tufts Clinical and Translational Science Institute N3C Supplement); University of Massachusetts Medical School Worcester (UL1TR001453: University of Massachusetts Center for Clinical and Translational Science); University of Alabama at Birmingham (UL1TR003096: Center for Clinical and Translational Science); University of Arkansas for Medical Sciences

(UL1TR003107: UAMS Translational Research Institute); The University of Chicago (UL1TR002389: ITM 2.0: Advancing Translational Science in Metropolitan Chicago); University of Colorado Denver (UL1TR002535-03S2: CCTSI Participation in the National COVID Cohort Collaborative N3C); University of Illinois at Chicago (UL1TR002003: Clinical and Translational Science Award); The University of Iowa (UL1TR002537: The University of Iowa Clinical and Translational Science Award); University of Kentucky (UL1TR001998-04S1: Kentucky Center for Clinical and Translational Science); University of Miami (UL1TR002736: Miami Clinical and Translational Science Institute); The University of Michigan at Ann Arbor (UL1TR002240: Michigan Institute for Clinical and Health Research); University of Minnesota (UL1TR002494: University of Minnesota Clinical and Translational Science Institute); University of Nebraska Lincoln (U54GM115458: University of Nebraska Center for Clinical & Translational Research); University of North Carolina at Chapel Hill (UL1TR002489: ICEES+ COVID-19 Open Infrastructure to Democratize and Accelerate Cross-Institutional Clinical Data Sharing and Research); University of Southern California (UL1TR001855: Southern California Clinical and Translational Institute); The University of Texas Medical Branch at Galveston (UL1TR001439: UTMB Clinical and Translational Science Award); The University of Utah (UL1TR002538-03S3: Infrastructure Support for Participation in the N3C Data Repository); University of Washington (UL1TR002319: Institute of Translational Health Sciences); University of Wisconsin-Madison (UL1TR002373: Institutional Clinical AND Translational Science Award); University of Virginia (UL1TR003015-02S2: Provision of Clinical Data to Support a Nationwide COVID-19 Cohort Collaborative); Virginia Commonwealth University (UL1TR002649-03S3: N3C & All of Us Research Program Collaborative Project); Wake Forest University Health Sciences (UL1TR001420: Wake Forest Clinical and Translational Science Award); Washington University in St. Louis (UL1TR002345: Washington University Institute of Clinical Translational Sciences); West Virginia University (U54GM104942: West Virginia Clinical and Translational Science Institute).

## Contributions

Contributions are organized according to contribution roles as follow:

### Data curation

Tellen D. Bennett, Richard A. Moffitt, Adit Anand, Zhenglong Qian, Katie Rebecca Bradwell, Davera Gabriel, Andrew T. Girvin, Stephanie S. Hong, Hunter Jimenez, Ramakanth Kavuluru, Kristin Kostka, Harold P. Lehmann, Amin Manna, Emily R. Pfaff, Nabeel Qureshi, Seth Russell, Peter E. DeWitt, Yun Jae Yoo, Richard L. Zhu, Ken R. Gersing, and Christopher G. Chute. These authors all had access to the limited dataset required for data analysis presented herein.

### Data integration

Richard A. Moffitt, Janos G. Hajagos, Benjamin Amor, Adit Anand, Mark M. Bissell, Katie Rebecca Bradwell, Davera Gabriel, Andrew T. Girvin, Stephanie S. Hong, Hunter Jimenez, Kristin Kostka, Amin Manna, Matvey B. Palchuk, Nabeel Qureshi, Seth Russell, Richard L. Zhu, Ken R. Gersing, Christopher G. Chute;

### Data quality assurance

Tellen D. Bennett, Richard A. Moffitt, Janos G. Hajagos, Adit Anand, Mark M. Bissell, Katie Rebecca Bradwell, Davera Gabriel, Andrew T. Girvin, Stephanie S. Hong, Hunter Jimenez, Kristin Kostka, Harold P. Lehmann, Eli Levitt, Amin Manna, Michele Morris, Matvey B. Palchuk, Emily R. Pfaff, Nabeel Qureshi, Seth Russell, Jacob T. Wooldridge, Yun Jae Yoo, Xiaohan Tanner Zhang, Richard L. Zhu, Joel H. Saltz, Ken R. Gersing, and Christopher G. Chute.

### Data visualization

Richard A. Moffitt, Adit Anand, Zhenglong Qian, Carolyn Bremer, James Brian Byrd, Alina Denham, Andrew T. Girvin, Elaine L. Hill, Hunter Jimenez, Amin Manna, Julie A. McMurry, Seth Russell, Peter E. DeWitt, Tellen D. Bennett, Yun Jae Yoo, Ken R. Gersing, and Melissa A. Haendel.

### Manuscript review and editing

Tellen D. Bennett, Richard A. Moffitt, Janos G. Hajagos, James Brian Byrd, Alina Denham, Brian T. Garibaldi, Andrew T. Girvin, Justin Guinney, Elaine L. Hill, Ramakanth Kavuluru, Eli Levitt, Sandeep K. Mallipattu, Julie A. McMurry, Emily R. Pfaff, Seth Russell, Heidi Spratt, Christopher P. Austin, Joel H. Saltz, Melissa A. Haendel, and Christopher G. Chute.

### Clinical subject matter expertise

Tellen D. Bennett, James Brian Byrd, Davera Gabriel, Brian T. Garibaldi, Eli Levitt, Sandeep K. Mallipattu, John Muschelli, Jacob T. Wooldridge, Xiaohan Tanner Zhang, Christopher P. Austin, Joel H. Saltz, Ken R. Gersing, and Christopher G. Chute.

### Manuscript drafting

Tellen D. Bennett, Richard A. Moffitt, James Brian Byrd, Brian T. Garibaldi, Andrew T. Girvin, Eli Levitt, Sandeep K. Mallipattu, Julie A. McMurry, Emily R. Pfaff, Heidi Spratt, Joel H. Saltz, Christopher

G. Chute, and Melissa A. Haendel.

### Project management

Richard A. Moffitt, Tellen D. Bennett, Davera Gabriel, Julie A. McMurry, Andrew J. Neumann, Nabeel Qureshi, Anita Walden, Christopher P. Austin, Ken R. Gersing, and Melissa A. Haendel.

### Funding acquisition

Julie A. McMurry, Nabeel Qureshi, Anita Walden, Christopher P. Austin, Ken R. Gersing, Melissa A. Haendel, and Christopher G. Chute.

### Database / information systems admin

Janos G. Hajagos, Andrew T. Girvin, Hunter Jimenez, Amin Manna, Julie

A. McMurry, Andrew J. Neumann, Anita Walden, Andrew E. Williams, and Ken R. Gersing.

### Clinical data model expertise

Tellen D. Bennett, Davera Gabriel, Ramakanth Kavuluru, Kristin Kostka, Harold P. Lehmann, Eli Levitt, Michele Morris, Emily R. Pfaff, Xiaohan Tanner Zhang, Richard L. Zhu, Joel H. Saltz, Ken R. Gersing; N3C Phenotype definition: Kristin Kostka, Michele Morris, Matvey B. Palchuk, Emily R. Pfaff, Andrew E. Williams, Ken R. Gersing, and Christopher G. Chute.

### Regulatory management and governance

Julie A. McMurry, Andrew J. Neumann, Anita Walden, Ken R. Gersing, Melissa A. Haendel, and Christopher G. Chute.

## Data Sharing

The N3C Data Enclave (covid.cd2h.org/enclave) houses fully reproducible, transparent, and broadly available limited and de-identified datasets (HIPAA definitions: https://www.hhs.gov/hipaa/for-professionals/privacy/special-topics/de-identification/index.html). Data is accessible by investigators at institutions that have signed a Data Use Agreement with NIH who have taken human subjects and security training and attest to the N3C User Code of Conduct. Investigators wishing to access the limited dataset must also supply an institutional IRB protocol. All requests for data access are reviewed by the NIH Data Access Committee. A full description of the N3C Enclave governance has been published;^3^ information about how to apply for access is available on the NCATS website: https://ncats.nih.gov/n3c/about/applying-for-access. Reviewers and health authorities will be given access permission and guidance to aid reproducibility and outcomes assessment. A Frequently Asked Questions about the data and access has been created at; https://ncats.nih.gov/n3c/about/program-faq

The data model is OMOP 5.3.1, specifications are posted at: https://ncats.nih.gov/files/OMOP_CDM_COVID.pdf

The latest version of the N3C Covid-19 Phenotype is always available at: https://github.com/National-COVID-Cohort-Collaborative/Phenotype_Data_Acquisition

Governance documents, codesets, code, and other N3C resources are available within the project Github repositories and/or in Zenodo for archival purposes:

https://github.com/National-COVID-Cohort-Collaborative

https://zenodo.org/communities/cd2h-covid/

Information on the source Common Data Models is available at: OHDSI: https://ohdsi.org/

PCORNet: https://pcornet.org/

ACT: https://www.dbmi.pitt.edu/node/53983

TriNetX: https://trinetx.com/

Other referenced resources are available at:

COVID-19 Map - Johns Hopkins Coronavirus Resource Center https://coronavirus.jhu.edu/map.html

Institutional Development Award Program Infrastructure for Clinical and Translational Research (IDeA-CTR) https://www.nigms.nih.gov/Research/DRCB/IDeA/Pages/IDeA-CTR.aspx

xgboost https://github.com/dmlc/xgboost

## Footnotes

[a]. https://coronavirus.jhu.edu/map

[b]. https://www.nigms.nih.gov/Research/DRCB/IDeA/Pages/IDeA-CTR.aspx

[c]. https://github.com/National-COVID-Cohort-Collaborative/Phenotype_Data_Acquisition

