## Supplemental Materials for "The National COVID Cohort Collaborative: Clinical Characterization and Early Severity Prediction"

**Supplemental Methods**	**1**

**Supplemental Tables**	**3**

Supplemental Table 2: Input Variables for Machine Learning	4

Supplemental Table 3a: N3C Cohort by Source Data Model	5

Supplemental Table 4: Multivariable Logistic Regression Models for Poor Outcome	6

Supplemental Table 5: Antimicrobials and Immunomodulation, Respiratory, Cardiovascular, and Renal Organ System Support for Hospitalized Patients, by Severity Group	7

Supplemental Table 5a: Antimicrobials and Immunomodulation	7

Supplemental Table 5b: Respiratory, Cardiovascular, and Renal Organ System Support for Hospitalized Patients, by Severity Group	7

Supplemental Table 6: Machine Learning Model Performance Metrics	8

**Supplemental Figures**	**9**

Supplemental Figure 1: Cohort Construction	9

Supplemental Figure 2: Trajectories of Additional Laboratory Tests During a Hospital Encounter	10

Supplemental Figure 3a: Heatmap showing Nadir, Average, and Peak Values of Vital signs and Body Size Metrics, by Severity Group	11

Supplemental Figure 3b: Heatmap showing Nadir, Average, and Peak Values of Laboratory Test Values, by Severity Group	12

Supplemental Figure 4: Relatively Few Patients have Harmonized Blood Type	13

### Supplemental Methods

*N3C Architecture, Data Integration and Harmonization Pipeline*

The N3C is hosted in a cloud-based, FedRAMP Moderate secure enclave^1^ managed by the National Center for Advancing Translational Sciences (NCATS). The N3C Enclave contains l1 Foundry, a data science platform that enables complex and reproducible analysis using a variety of open-source languages (e.g. Python^2^, R^3^, SQL^4^, Java^5^, and also point-and-click and dashboard-style tools. Foundry uses Apache Spark^6^ to support distributed operations on very large data sources.

Contributing sites submit a Health Insurance Portability and Accountability Act (HIPAA)-defined limited data set in one of four common data models (CDMs: PCORNet^7^ , the Observational Medical Outcomes Partnership [OMOP]^8^, ACT/i2b2^9^, or TriNetX^10^. Sites send updated data payloads approximately weekly. N3C harmonizes site data into OMOP version 5.3.1 in partnership with subject matter experts from each CDM community. In this process, all data (e.g. laboratory measurements, clinical observations such as vital signs, medications, and clinical conditions, are harmonized and mapped to the OMOP vocabulary^8^. Site data that pass a robust data quality assessment pipeline are integrated into the “release” set for use by the community. Details about the data transfer, harmonization, quality, and integration processes have been reported^11^.

*Identifying Hospital Encounters, Comorbidities, Medications, Mechanical Ventilation, Vital Signs, and Laboratory Tests*

We defined a single index encounter for each laboratory-confirmed positive patient by selecting encounters that start up to 30 days before or 7 days after the positive test result, or a positive test result occurs during the visit. When multiple encounters met these criteria, we broke ties by preferentially selecting the encounter in which the most severe outcome was observed, then the longest visit, and finally the most recent visit.

We reconstructed hospital encounters from component “visits” (e.g. a radiology study and a surgical procedure recorded as separate visits) using an algorithm that will be made available to all N3C users. We built hospital encounters from recorded OMOP visits by first filtering to Inpatient (9201), Inpatient Hospital (8717), Intensive Care (32037), Emergency Room (ER) and Inpatient (262), or Inpatient Critical Care Facility (581379) visits of any duration, ER visits (9203) spanning at least 2 calendar days, or Outpatient Visits (9202) spanning exactly two calendar days. These visits were then merged such that any visits with overlapping calendar days would end up in the same hospital stay. Finally, all merged visits that did not contain at least one inpatient or ER visit were unmerged. This process results in combined hospital stays that are separated by a period of at least one calendar day. Finally, visits of any type that occur during a combined hospital stay are added to the hospital stay.

All OMOP concept sets developed for this manuscript are freely available on the platform, versioned, and include attributed input from both informatics and clinical subject matter experts. None of the 4 CDMs support admission, discharge, and transfer (ADT) tables, which complicates analyses of hospital encounters.

We defined comorbidities based on the updated^12^ Charlson Comorbidity Index as implemented in the ‘icd’ R package^13^. Unless otherwise noted, we identified medications using the WHO anatomical therapeutic chemical (ATC) definitions.^14^ We built an invasive ventilation concept set from standardized terminology codes (International Classification of Diseases [ICD] and Systematized Nomenclature of Medicine [SNOMED]) included in the OMOP CDM. Among hospitalized patients, we assessed serial measurements of heart rate (HR), respiratory rate (RR), temperature, systolic and diastolic blood pressure (SBP and DBP, respectively), pulse oximetry (SpO_2_), and a variety of laboratory tests.

*Software*

We used reproducible pipelines in SQL, R, and Python to conduct all analyses. Our pipelines relied on the *SparkR**^15^* and *pyspark**^16^* interfaces to Apache Spark^6^. We built machine learning models using Python’s ‘scikit-learn’^17^ package and visualizations using R’s ‘ggplot2’^18^, ‘ggalluvial’^19^, and ‘ggnewscale’ packages^20^ and Python’s Matplotlib package.^21^

*Outcome Prediction Supplemental Methods*

Categorical variables were converted to k-1 dummy variables using Pandas’ get_dummies (one-hot encoding). For logistic regression and support vector machines, numeric variables were centered to mean zero with unit variance using scikit-learn’s StandardScaler. Optimal model specific hyperparameters were selected with a grid search performed using scikit-learn’s GridSearchCV using 5-fold cross validation on the training set with AUROC as the scoring metric. Each grid search included multiple iterations with categorical settings such as solver and with first coarse settings for numeric parameters following a logarithmic scale followed by more specific settings around the values found to perform best.

### Supplemental Tables

| **Variable, n(%) unless otherwise indicated** | **All** | **Lab-**  **confirmed Positive** | **Lab-**  **confirmed Negative** | **Suspected Positive** | **No Test for SARS- CoV-2** |
| --- | --- | --- | --- | --- | --- |
| **Age, mean (SD)** | 43.2 (22.9) n=192,5699 | 41.4 (20.4) n=199,935 | 44.2 (22.6) n=1,339,933 | 39.2 (26.2) n=174,831 | 41.6 (23.1) n=211,000 |
| **Sex** |  |  |  |  |  |
| **Female** | 1,074,141 | 106,316 | 750,606 | 96,073 | 121,146 |
| **Male** | 851,007 | 93,607 | 588,750 | 78,904 | 89,746 |
| **Other*** | 1,378 | 139 | 815 | 205 | 219 |
| **Race** |  |  |  |  |  |
| **White** | 1,251,401 | 104,491 | 898,340 | 108,992 | 139,578 |
| **Black or African- American** | 301,994 | 36,243 | 198,569 | 35,532 | 31,650 |
| **Native Hawaiian or Pacific Islander** | 3,034 | 459 | 2,000 | 265 | 310 |
| **Asian** | 48,897 | 4,690 | 35,106 | 4,188 | 4,913 |
| **Other** | 20,626 | 2,363 | 1,3813 | 1,405 | 3045 |
| **Missing/Unknown** | 300,574 | 51,816 | 192,343 | 24,800 | 31,615 |
| **Ethnicity** |  |  |  |  |  |
| **Hispanic** | 156,401 | 34,657 | 93,137 | 16,668 | 11,939 |
| **Non-Hispanic** | 1,498,261 | 130,297 | 1,072,760 | 126,982 | 168,222 |
| **Missing/Unknown** | 300,574 | 51,816 | 192,343 | 24,800 | 31,615 |
| **Insurance Payer** |  |  |  |  |  |
| **Medicare** | 118,381 | 7,416 | 87,102 | 9,924 | 13,939 |
| **Commercial** | 212,527 | 17,247 | 137,233 | 15,437 | 42,610 |
| **Medicaid** | 106,558 | 9,532 | 66,677 | 12,436 | 17,913 |
| **Other** | 1,783,181 | 186,593 | 1,252,174 | 149,335 | 195,079 |

**Supplemental Table 1: N3C Cohort Characteristics**

**Supplemental Table 1**: This table shows demographic characteristics and insurance payer for the overall N3C cohort, stratified by the N3C phenotype groups (publicly available on GitHub^22^). SARS-CoV-2 = severe acute respiratory syndrome associated with coronavirus-2. *Other includes non-binary, no matching concept, and no information.

##### Supplemental Table 2: Input Variables for Machine Learning

| This table shows the 42 categories of 64 input variables for the machine learning models. The worst value for each variable on the first calendar day of hospital admission was used. We defined the worst value as the lowest value for diastolic blood pressure, hemoglobin, pH, platelet count, SpO2, and systolic blood pressure. For the remainder, we used the highest value. NTproBNP = N-Terminal-prohormone B-type Natriuretic Peptide. *(White, Black or African-American, Native Hawaiian or Pacific Islander, Other, or Missing/Unknown) | **Variable (units)** | **%present** | **Imputation Strategy** |
| --- | --- | --- | --- |
|  | Age at visit start (years) | 100.0% | None |
|  | Sex (Female, Male, or Other) | 100.0% | Missing values filled with ‘Other’ |
|  | White blood cell count (x10E3/uL) | 94.2% | Median |
|  | Platelet count (x10E3/uL) | 94.1% | Median |
|  | Hemoglobin (g/dL) | 93.2% | Median |
|  | Creatinine (mg/dL) | 92.9% | Median |
|  | Sodium (mmol/L) | 92.8% | Median |
|  | BUN (mg/dL) | 92.7% | Median |
|  | Chloride (mmol/L) | 92.7% | Median |
|  | Potassium (mmol/L) | 92.5% | Median |
|  | Glucose (mg/dL) | 92.1% | Median |
|  | Ethnicity (Hispanic, Not Hispanic, or Missing/Unknown) | 88.8% | Missing values filled with ‘Missing/Unknown’ |
|  | Aspartate Aminotransferase (AST/SGOT, IU/L) | 83.4% | Median |
|  | Bilirubin (total, mg/dL) | 82.9% | Median |
|  | Race* | 76.7% | Missing values filled with ‘Missing/Unknown’ |
|  | Alanine Aminotransferase (ALT/SGPT, IU/L) | 75.7% | Median |
|  | Absolute Lymphocyte count (x10E3/uL) | 74.6% | Median |
|  | Body Weight (kg) | 73.3% | Median |
|  | Absolute Neutrophil count (x10E3/uL) | 70.2% | Median |
|  | Diastolic blood pressure (DBP) | 65.9% | Median |
|  | Systolic blood pressure (SBP) | 65.9% | Median |
|  | Albumin (g/dL) | 57.5% | Median |
|  | Oxygen saturation (SpO2) | 53.5% | Median |
|  | Ferritin (ng/mL) | 49.3% | Male and missing: 150; Female and missing: 75 |
|  | Respiratory Rate | 49.3% | Median |
|  | C-reactive protein (CRP, mg/L) | 49.1% | Missing values filled with 10 |
|  | Charlson Cancer | 48.6% | FALSE |
|  | Charlson Congestive heart failure (CHF) | 48.6% | FALSE |
|  | Charlson Dementia | 48.6% | FALSE |
|  | Charlson Diabetes Mellitus | 48.6% | FALSE |
|  | Charlson Diabetes Mellitus with complications | 48.6% | FALSE |
|  | Charlson HIV | 48.6% | FALSE |
|  | Charlson Liver disease (mild) | 48.6% | FALSE |
|  | Charlson Liver disease (severe) | 48.6% | FALSE |
|  | Charlson Metastases | 48.6% | FALSE |
|  | Charlson Myocardial Infarction | 48.6% | FALSE |
|  | Charlson Hemiplegia or paralysis | 48.6% | FALSE |
|  | Charlson Peptic ulcer disease | 48.6% | FALSE |
|  | Charlson Pulmonary disease | 48.6% | FALSE |
|  | Charlson Peripheral vascular disease | 48.6% | FALSE |
|  | Charlson Comorbidity Index, Q score | 48.6% | Missing values filled with 0 |
|  | Charlson Renal disease | 48.6% | FALSE |
|  | Charlson Rheumatologic Disease | 48.6% | FALSE |
|  | Charlson Stroke | 48.6% | FALSE |
|  | Body mass index (BMI, kg/m2) | 48.3% | Median |
|  | Temperature | 46.1% | Median |
|  | Lactate (mM/L) | 45.5% | Missing values filled with 13.5 |
|  | D-Dimer (mg/L FEU) | 43.7% | Median |
|  | Troponin all types (ng/mL) | 43.2% | Median |
|  | Heart rate | 34.1% | Median |
|  | Bilirubin (conjugated/direct, mg/dL) | 27.0% | Median |
|  | pH | 26.4% | Median |
|  | Procalcitonin (ng/mL) | 24.9% | Missing values filled with 0.02 |
|  | Hemoglobin-glycosylated (A1C, %) | 20.2% | Median |
|  | Erythrocyte Sedimentation Rate (mm/hr) | 19.6% | Missing values filled with 19 |
|  | NTproBNP (pg/mL) | 18.3% | Missing values filled with 125 |
|  | BNP (pg/mL) | 16.3% | Missing values filled with 100 |

##### Supplemental Table 3a: N3C Cohort by Source Data Model

This table shows the representation among N3C sites of each common data model (CDM). CDMs include the National Patient-Centered Clinical Research Network (PCORNet),^7^ the Observational Health Data Sciences and Informatics (OHDSI) network,^23^ the Accrual to Clinical Trials (ACT) network,^9^ and TriNetX^10^. This table includes a total of 36 sites. Two sites are dropped prior to analysis due to missing date data (see Supplemental Figure 1).

| **Data model** | **# N3C Sites** | **# Patients represented** |
| --- | --- | --- |
| **OMOP** | **6** | 305,376 |
| **PCORnet** | **12** | 1,036,073 |
| **i2b2/ACT** | **6** | 359,920 |
| **TriNetX** | **10** | 444,690 |

**Supplemental Table 3b: Variables Supported by Source Data Models[1]**

S = supported, NS = not supported. **[1]** Variables *supported* by a data model may not be *required* by that model to conform to the model’s specification. Thus, some systematic missingness may be at the site level rather than the model level. **[2]** Many of the items marked “not supported” for ACT can technically be stored in the i2b2 data model, which underlies ACT; however, they are not supported by the ACT ontology at this time, and are not harmonized by N3C. **[3]** All models support both quantitative and qualitative lab results; however, many sites only map a subset of their qualitative lab results to the model’s vocabulary. *A small set of vitals are defined by the model; additional vital data can optionally be modelled as “observations”

|  | **OMOP** | **PCORnet** | **ACT[2]** | **TriNetX** |
| --- | --- | --- | --- | --- |
| Patient Demographics | S | S | S | S |
| Visit (encounter) details | S | S | S | S |
| Discharge disposition | S | S | NS | NS |
| Diagnoses | S | S | S | S |
| Medications | S | S | S | S |
| Laboratory results[3] | S | S | S | S |
| Procedures | S | S | S | S |
| Vital signs | S | S* | NS | S |
| Location of patient residence (ZIP code-level) | S | S | NS | S |
| Death | S, date required | S, date not required | S, date not required | S, date required |
| Admission - Discharge - Transfer transactions | NS | NS | NS | NS |
| Insurance | S | S | NS | NS |

##### Supplemental Table 4: Multivariable Logistic Regression Models for Poor Outcome

This table shows odds ratios (ORs) and 95% confidence intervals (CIs) for 2 multivariable logistic regression models, one with missing/unknown as a category when relevant and one with complete cases only. LCL = lower confidence limit, lower bound of 95% CI. UCL = upper confidence limit, upper bound of 95% CI. See Results for details. These models were built after the prediction models and are for inference only.

|  |  |  | **Missing encoded** | |  | **Missing cases dropped** | |
| --- | --- | --- | --- | --- | --- | --- | --- |
|  |  |  | **OR** | **p-value** |  | **OR** | **p-value** |
| **Age** | per year |  | 1.03 | p < 0.0001 |  | 1.03 | p < 0.0001 |
| **Comorbidities** | Diabetes mellitus |  | 1.05 | p = 0.2106 |  | 0.85 | p = 0.1557 |
|  | Liver disease |  | 1.20 | p = 0.0010 |  | 1.07 | p = 0.6915 |
|  | Cancer |  | 0.96 | p = 0.3922 |  | 0.85 | p = 0.2403 |
|  | Pulmonary |  | 0.93 | p = 0.0886 |  | 0.91 | p = 0.4259 |
|  | Renal |  | 1.06 | p = 0.2053 |  | 0.91 | p = 0.4938 |
|  | Congestive Heart Failure |  | 1.07 | p = 0.1226 |  | 0.88 | p = 0.3431 |
|  | Rheumatic Disease |  | 0.83 | p = 0.0151 |  | 0.93 | p = 0.7210 |
|  | Dementia |  | 1.26 | p < 0.0001 |  | 0.80 | p = 0.2761 |
|  | none of the above |  | 1.00 | ref. |  | 1.00 | ref. |
| **Gender** | Male |  | 1.60 | p < 0.0001 |  | 1.70 | p < 0.0001 |
|  | Female |  | 1.00 | ref. |  | 1.00 | ref. |
| **Ethnicity** | Hispanic or Latino |  | 1.04 | p = 0.4663 |  | 1.04 | p = 0.8381 |
|  | Not Hispanic or Latino |  | 1.00 | ref. |  | 1.00 | ref. |
|  | *unknown* |  | 1.14 | p = 0.0057 |  |  |  |
| **Race** | Black or African-American |  | 1.12 | p = 0.0011 |  | 1.21 | p = 0.0930 |
|  | Asian |  | 1.33 | p = 0.0011 |  | 2.36 | p = 0.0017 |
|  | Other |  | 1.25 | p = 0.0477 |  | 1.22 | p = 0.4255 |
|  | White |  | 1.00 | ref. |  | 1.00 | ref. |
|  | *unknown* |  | 1.19 | p = 0.0005 |  |  |  |
| **BMI** | over 30 |  | 1.36 | p < 0.0001 |  | 1.41 | p = 0.0008 |
|  | 30 or under |  | 1.00 | ref. |  | 1.00 | ref. |
|  | *unknown* |  | 1.23 | p < 0.0001 |  |  |  |
| **Blood type** | A |  | 0.90 | p = 0.2660 |  | 0.93 | p = 0.4910 |
|  | B |  | 0.97 | p = 0.7884 |  | 1.12 | p = 0.4405 |
|  | AB |  | 0.55 | p = 0.0256 |  | 0.53 | p = 0.0353 |
|  | O |  | 1.00 | ref. |  | 1.00 | ref. |
|  | *unknown* |  | 0.37 | p < 0.0001 |  |  |  |
| **Rh factor** | negative |  | 0.94 | p = 0.6621 |  | 1.11 | p = 0.5477 |
|  | positive |  | 1.00 | ref. |  | 1.00 | ref. |

##### Supplemental Table 5: Antimicrobials and Immunomodulation, Respiratory, Cardiovascular, and Renal Organ System Support for Hospitalized Patients, by Severity Group

###### Supplemental Table 5a: Antimicrobials and Immunomodulation

This table shows the percent of patients in each category who received each medication type. ED = Emergency Department. WHO = World Health Organization. ECMO = extracorporeal membrane oxygenation. LOS = length of stay. We stratified patients using the Clinical Progression Scale (CPS) established by the World Health Organization (WHO) for COVID-19 clinical research.^24^ Severity assigned by patient-specific encounter maximum severity.

###### Supplemental Table 5b: Respiratory, Cardiovascular, and Renal Organ System Support for Hospitalized Patients, by Severity Group

This table shows the percent of patients in each category who received each treatment type. ED = Emergency Department. WHO = World Health Organization. ECMO = extracorporeal membrane oxygenation. LOS = length of stay. We stratified patients using the Clinical Progression Scale (CPS) established by the World Health Organization (WHO) for COVID-19 clinical research.^24^ Severity assigned by patient-specific encounter maximum severity. CRRT = Continuous Renal Replacement Therapy. HD = hemodialysis.

| **A** |  |  |  |  | **B** |  |  |  |
| --- | --- | --- | --- | --- | --- | --- | --- | --- |
|  | **Moderate**  **Hospitalized without invasive ventilation**  **WHO**  **Severity 4-6** | **Severe**  **Hospitalized with invasive ventilation or ECMO**  **WHO**  **Severity 7-9** | **Hospital Mortality or Discharge to Hospice**  **WHO Severity 10** |  |  | **Moderate**  **Hospitalized without invasive ventilation**  **WHO**  **Severity 4-6** | **Severe**  **Hospitalized with invasive ventilation or ECMO**  **WHO**  **Severity 7-9** | **Hospital Mortality or Discharge to Hospice**  **WHO Severity 10** |
| **Antimicrobials** |  |  |  |  | **Respiratory Support** | |  |  |
| Remdesivir | 15.31% | 25.13% | 20.08% |  | Inhaled Nitric Oxide | 0.00% | 0.04% | 0.08% |
| Lopinavir/Ritonavir | 0.36% | 1.94% | 1.09% |  | Epoprostenol | 0.11% | 7.06% | 6.52% |
| Hydroxychloroquine | 6.85% | 21.76% | 14.65% |  |  |  |  |  |
| Chloroquine | 0.04% | 0.61% | 0.19% |  | **Cardiovascular Support** | |  |  |
| Any Antibacterial | 54.22% | 89.93% | 78.70% |  | Amiodarone | 0.63% | 8.60% | 12.64% |
| Any Antiviral | 3.18% | 7.96% | 5.78% |  | Dopamine | 2.07% | 7.42% | 7.02% |
| Any Antifungal | 2.20% | 15.99% | 13.85% |  | Dobutamine | 0.17% | 3.48% | 2.86% |
|  |  |  |  |  | Epinephrine | 1.02% | 10.00% | 14.38% |
| **Immunomodulation** | |  |  |  | Esmolol | 0.30% | 2.69% | 1.72% |
| Dexamethasone | 9.45% | 13.66% | 9.43% |  | Isoproterenol | 0.02% | 0.07% | 0.08% |
| Prednisone | 7.76% | 16.42% | 10.15% |  | Milrinone | 0.04% | 1.11% | 0.61% |
| Methylprednisolone | 4.42% | 21.15% | 16.00% |  | Norepinephrine | 0.44% | 14.66% | 11.89% |
| Hydrocortisone | 1.00% | 14.34% | 18.31% |  | Phenylephrine | 1.04% | 12.87% | 10.20% |
| Any systemic steroid | 35.83% | 67.46% | 56.21% |  | Vasopressin | 1.04% | 12.87% | 10.20% |
| Anakinra | 0.05% | 0.04% | 0.13% |  | ECMO | 0.00% | 5.02% | 2.49% |
| Tocilizumab | 0.78% | 13.55% | 6.33% |  | CRRT or HD | 1.53% | 9.75% | 9.59% |

##### Supplemental Table 6: Machine Learning Model Performance Metrics

This table shows performance metrics for each machine learning model type over Inpatient stays ending between January 2020 and November 2020. Mar-May = March to May 2020. Jun-Oct = June to October 2020. AUROC = area under the receiver operator characteristic curve.

|  |  | **Random Forest** | **XGBoost** | **Support Vector Machines** | **Logistic Regression** | | |
| --- | --- | --- | --- | --- | --- | --- | --- |
|  |  |  |  |  | **None** | **L1** | **L2** |
| Balanced Accuracy | All | 68.2% | 71.1% | 62.6% | 66.0% | 65.3% | 66.2% |
|  | Jun-Oct | 65.4% | 68.3% | 61.2% | 64.0% | 63.4% | 64.0% |
|  | Mar-May | 70.1% | 73.0% | 63.8% | 67.6% | 66.9% | 67.8% |
| F1 | All | 52.4% | 57.0% | 40.4% | 47.7% | 46.2% | 47.9% |
|  | Jun-Oct | 45.3% | 50.0% | 36.2% | 41.7% | 40.5% | 41.7% |
|  | Mar-May | 56.9% | 61.8% | 43.4% | 51.7% | 50.4% | 52.2% |
| Precision | All | 80.3% | 74.7% | 82.4% | 72.4% | 72.0% | 72.1% |
|  | Jun-Oct | 71.3% | 64.8% | 74.0% | 62.3% | 61.9% | 61.7% |
|  | Mar-May | 87.2% | 83.1% | 89.5% | 79.4% | 79.0% | 79.6% |
| AUROC | All | 86.4% | 87.4% | 82.7% | 83.5% | 83.4% | 83.5% |
|  | Jun-Oct | 86.1% | 86.6% | 82.0% | 83.0% | 82.8% | 83.1% |
|  | Mar-May | 86.8% | 88.4% | 83.7% | 84.5% | 84.5% | 84.4% |
| Recall | All | 38.9% | 46.1% | 26.7% | 35.5% | 34.1% | 35.9% |
|  | Jun-Oct | 33.2% | 40.7% | 24.0% | 31.4% | 30.1% | 31.5% |
|  | Mar-May | 42.2% | 49.3% | 28.7% | 38.3% | 37.0% | 38.8% |

### Supplemental Figures

##### Supplemental Figure 1: Cohort Construction

This Sankey plot shows how the cohort accumulated from the N3C sites. The left vertical axis shows the number of patients (M = 1,000,000). Each site has a color. The width of the arrows corresponds to the number of patients from that site. Two sites did not submit sufficient date information for us to calculate the N3C computable phenotypes (publicly available on GitHub^22^), excluded as noted “FALSE” in the middle column. We then excluded a) sites who did not submit sufficient death and ventilation information and b) children < 18 years old (“removed from study”), and c) sites whose patients were overwhelmingly children. We show laboratory-confirmed positive as “PCR pos” in this plot due to limited space, but <5% of the patients at one site were included with SARS-CoV-2 antigen positivity. The remainder had positive SARS-CoV-2 polymerase chain reaction (PCR) tests. See Methods for details.

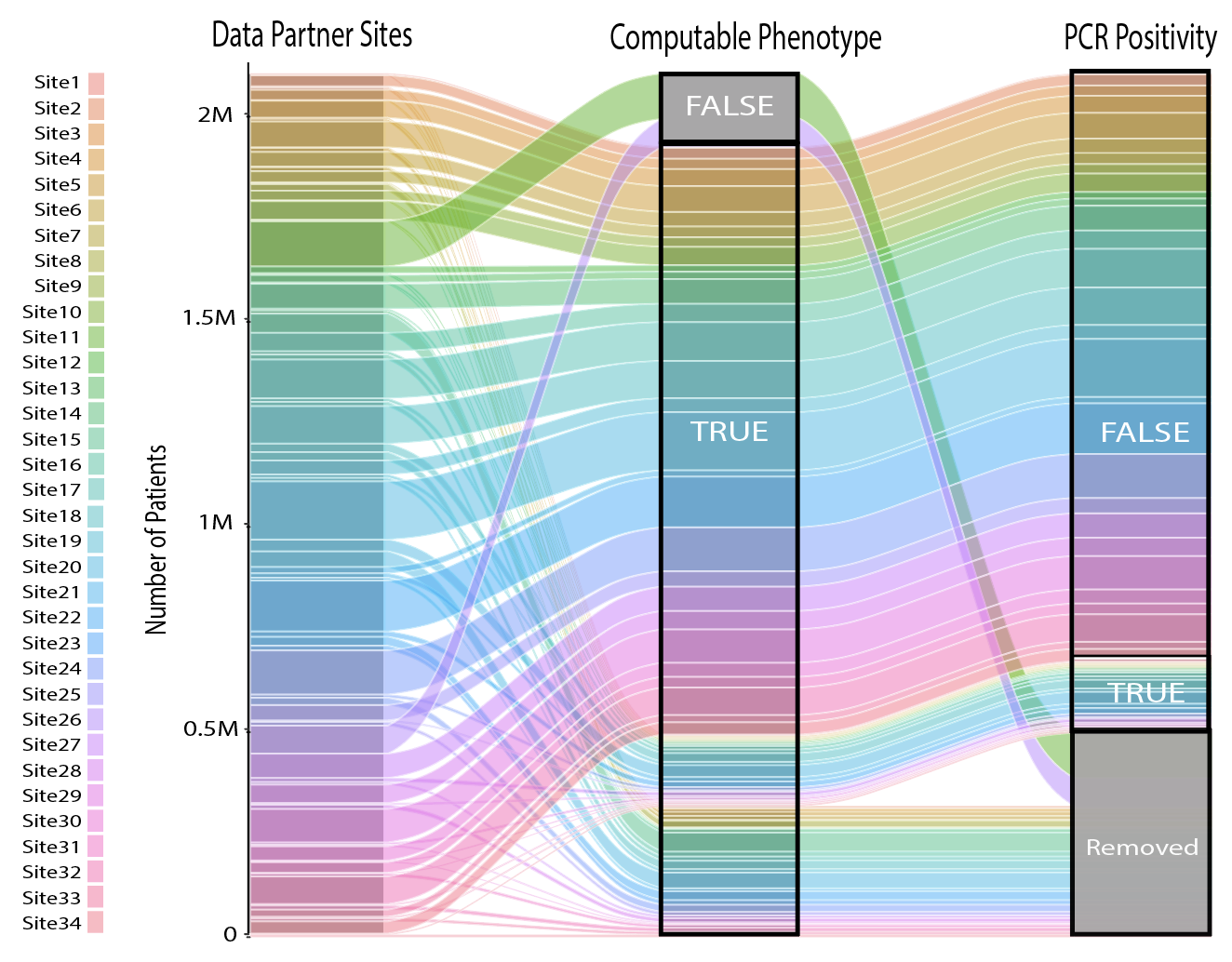

##### Supplemental Figure 2: Trajectories of Additional Laboratory Tests During a Hospital Encounter

This figure shows the median (line) and interquartile range (bars) of each laboratory test on each hospital day, stratified by patient maximum severity (hospital mortality or discharge to hospice [black], invasive ventilation or extracorporeal membrane oxygenation [red], hospitalized without any of those [yellow], or emergency department visit only [green], see Table 1). ALT = alanine aminotransferase. AST = aspartate aminotransferase. BUN = blood urea nitrogen. Sed. = sedimentation (erythrocyte sedimentation rate). IL-6 = interleukin-6. NTproBNP = N-Terminal-prohormone B-type Natriuretic Peptide.

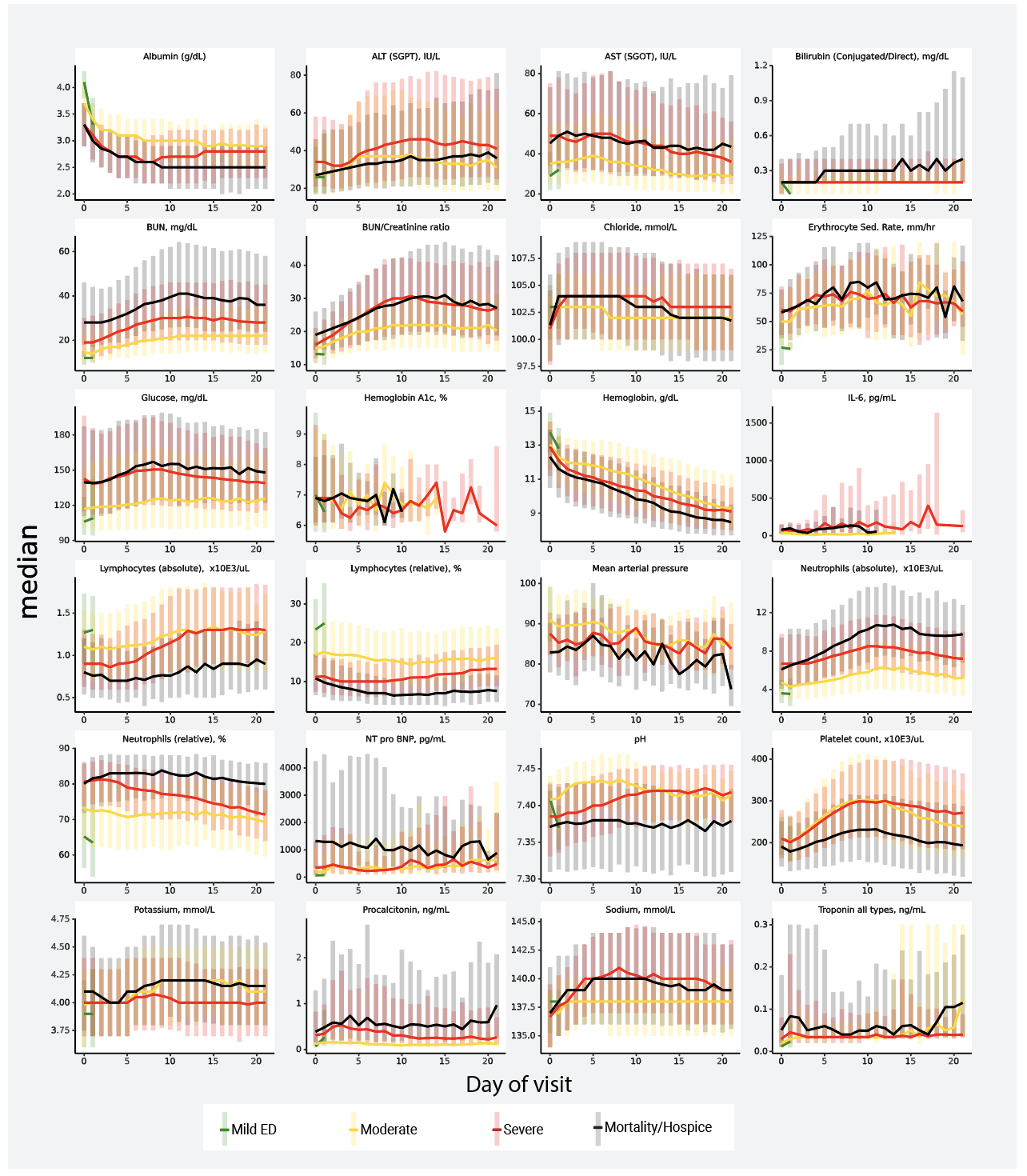

##### Supplemental Figure 3a: Heatmap showing Nadir, Average, and Peak Values of Vital signs and Body Size Metrics, by Severity Group

Values shown for each vital sign and body size metric for each severity group are multiples of the interquartile range (IQR) away from the median value. Circle diameter corresponds to the number of IQRs away from the median, with blue representing below the median and red representing above the median.

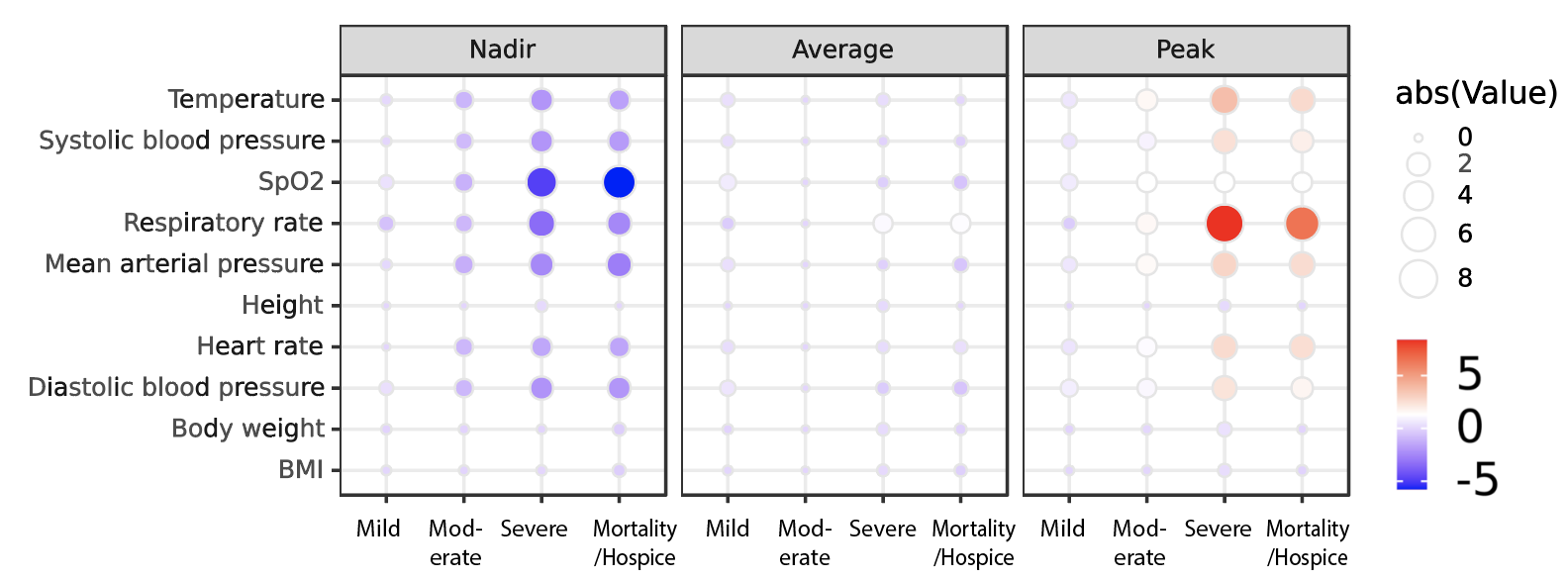

##### Supplemental Figure 3b: Heatmap showing Nadir, Average, and Peak Values of Laboratory Test Values, by Severity Group

Values shown for each laboratory test for each severity group are multiples of the interquartile range (IQR) away from the median value. Circle diameter corresponds to the number of IQRs away from the median, with blue representing below the median and red representing above the median. ALT = alanine aminotransferase. AST = aspartate aminotransferase. BUN = blood urea nitrogen. Sed. = sedimentation (erythrocyte sedimentation rate). IL-6 = interleukin-6. NTproBNP = N-Terminal-prohormone B-type Natriuretic Peptide.

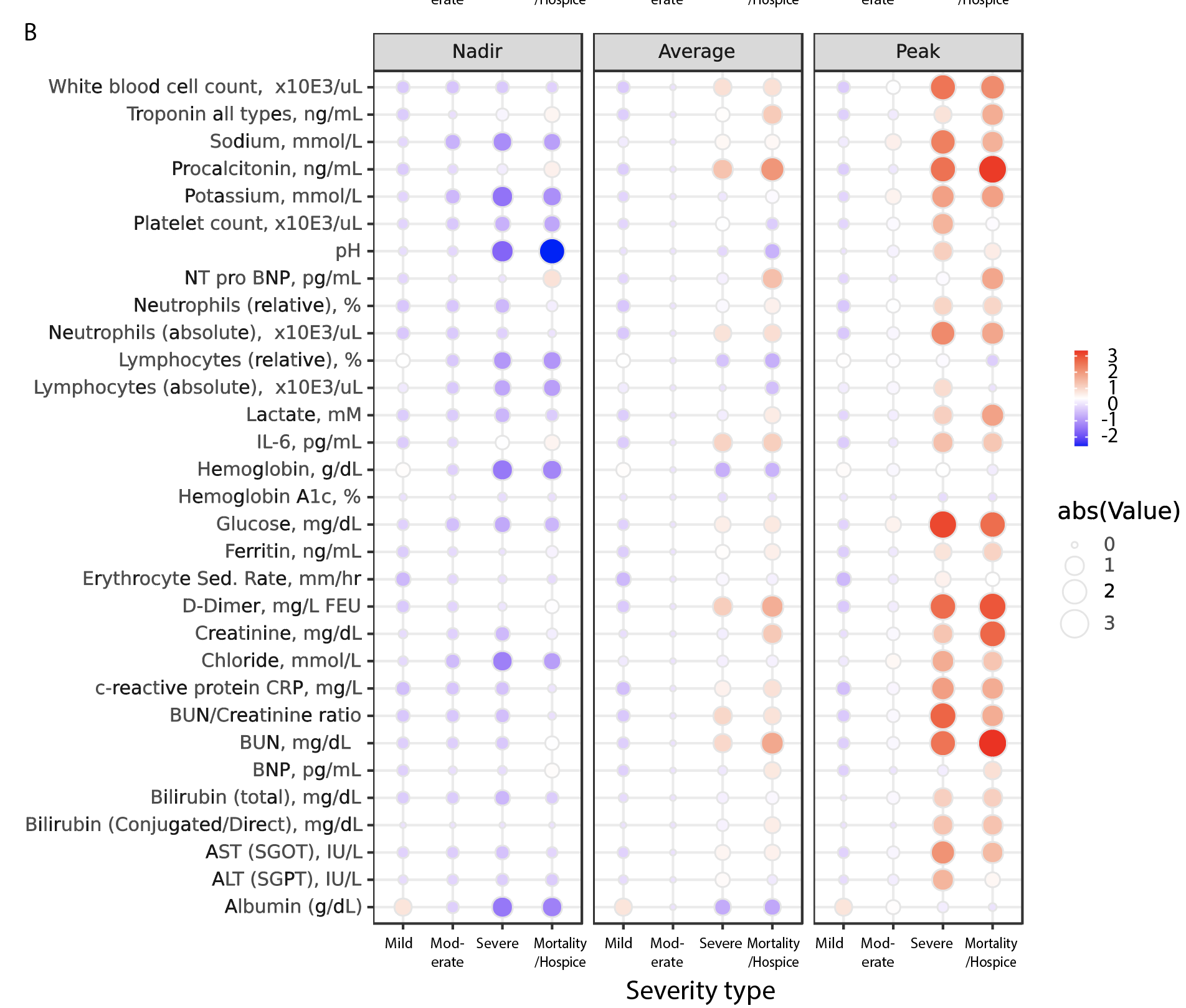

##### Supplemental Figure 4: Relatively Few Patients have Harmonized Blood Type

This sunburst plot^25^ is read from inside out. Each arc length corresponds to the proportion of that circle represented by that category. Composite patterns are shown as adjacent segments (inside to out), e.g. known blood type, type A, and positive Rh antigen. ED = Emergency Department. Neg = Rh negative. Pos = Rh positive. Stratification is by severity group.

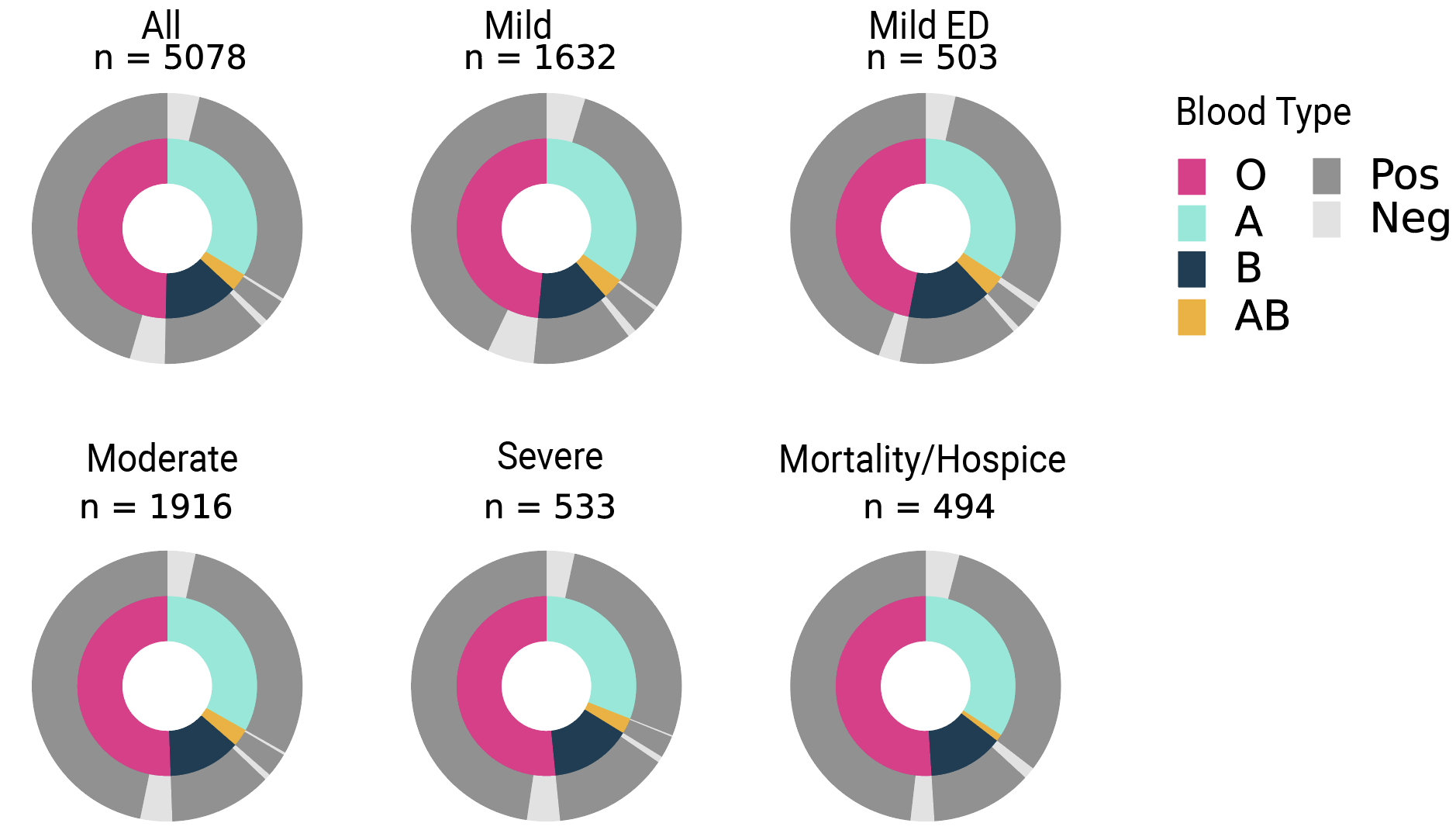

**Supplemental Figure 5a Title: Antimicrobial Treatments in Hospitalized Patients**

**Supplemental Figure 5a Legend:** This sunburst plot^25^ is read from inside to outside. Each arc length corresponds to the proportion of that circle represented by that category. Composite treatment regimens are shown as adjacent segments (inside to out, e.g. systemic antibiotic yes, azithromycin yes, hydroxychloroquine yes). Stratification is by severity group.

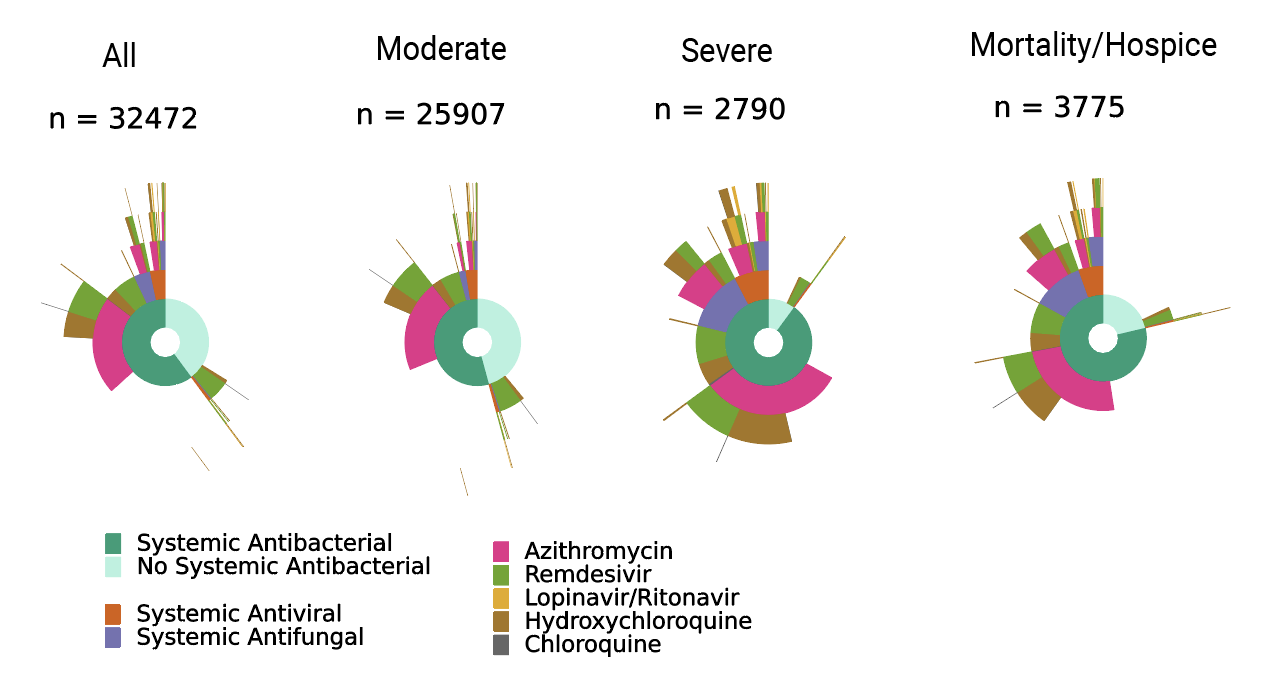

**Supplemental Figure 5b Title: Immunomodulatory Treatments in Hospitalized Patients**

**Supplemental Figure 5a Legend:** This sunburst plot is read from inside to outside.^25^ Each arc length corresponds to the proportion of that circle represented by that category. Composite treatment regimens are shown as adjacent segments (inside to out, e.g. systemic corticosteroid yes, dexamethasone yes, anakinra yes). Stratification is by severity group.

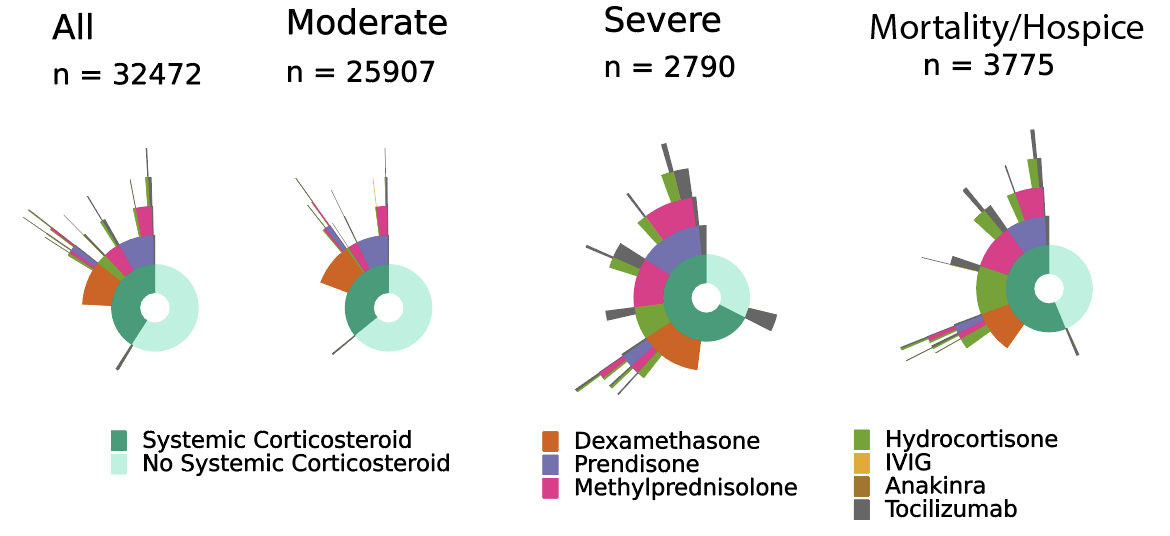

**Supplemental Figure 6 Title: Area Under the Receiver Operator Characteristic Curve (AUROC) for First-Day Machine Learning Models to Predict Subsequent Clinical Severity**

**Supplemental Figure 6 Legend**: AUC = AUROC. SVM = support vector machines. Logistic regression is shown with no penalization and L1 and L2 penalization. See Methods for details.

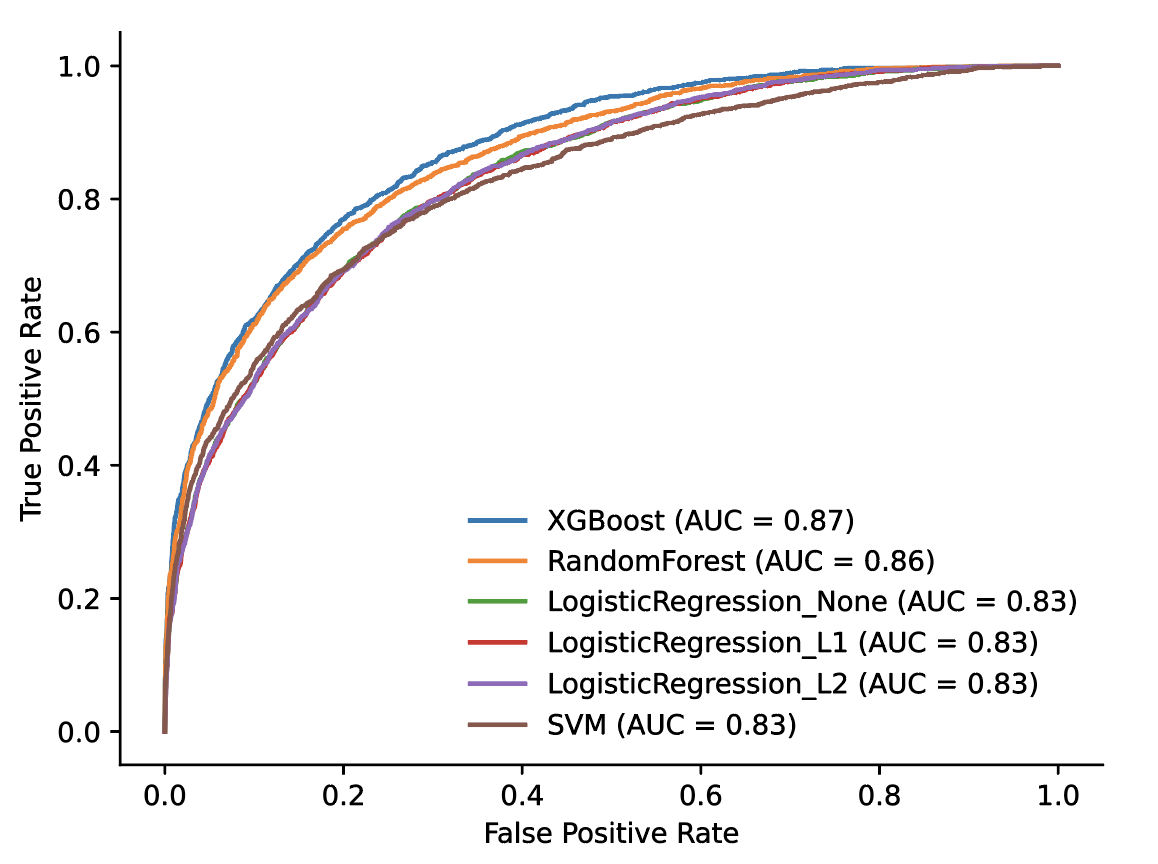
